# A systematic review with Bayesian modelling of the prevalence of neuropathic pain

**DOI:** 10.64898/2026.01.14.26344090

**Authors:** Peter R Kamerman, Taskeen Hoosen, Nkazimulo Mnguni, Prince C Chikezie

**Author notes:** **Corresponding author** Peter R Kamerman, Brain Function Research Group, Department of Physiology, School of Biomedical Sciences, Faculty of Health Sciences, University of the Witwatersrand, 7 York Road, Parktown, 2193, South Africa.

## Abstract

**Objective:** The prevalence of neuropathic pain has been estimated using imperfect screening tools, which can lead to inflated estimates. We performed a systematic review and meta-analysis of the prevalence of pain with neuropathic characteristics using Bayesian methods to correct prevalence estimates for imperfect screening tool sensitivity and specificity.

**Methods:** We searched major databases for national and regional epidemiological studies published between 1 January 2001 and 4 February 2025 that reported the prevalence of pain with neuropathic characteristics, as identified by the painDETECT, S-LANSS, or DN4-interview screening tools. In duplicate, reviewers extracted data on the number of individuals reported as having/not having pain with neuropathic characteristics. Corrected prevalence was estimated using Bayesian models that incorporated sensitivity and specificity distributions under four different priors [noninformative prior: Beta(1, 1), informative prior with a mode of 2.5%: Beta(1.398, 16.522), informative prior with a mode of 5%: Beta(2.063, 21.197), and informative prior with a mode of 10%: Beta(5.619, 42.5712)].

**Results:** From 1,251 records retrieved, 8 were extracted. While apparent prevalence point estimates ranged from 6.9% to 17.9%, the corrected Bayesian posterior point estimates ranged from 0.9% to 8.5%. The median (range) of the difference between the apparent point estimate for each study and the corresponding Bayesian posterior estimate for the non-informative prior was −8.4 percentage points (−12.6 to −5.8), reducing to −4.9 percentage points (−9.4 to −3.3) for the informative prior centered at 10% prevalence.

**Conclusion:** The use of imperfect screening tools may overestimate the prevalence of neuropathic pain.

## Introduction

Neuropathic pain is a type of pain caused by damage or disease of the somatosensory nervous system [1], and is associated with a significant health burden; more so than other causes of chronic pain [2, 3]. A systematic review of epidemiological studies estimated that between 6.9% and 10% of the general adult population has neuropathic pain [4]. However, the estimates of the prevalence of neuropathic pain in the general adult population are flawed because the instruments used to screen for the presence of neuropathic pain only assess for pains with neuropathic characteristics, which does not necessarily mean the pain is indeed neuropathic, but, more importantly, the instruments are not perfect tools. For example, screening tools such as the self-report Leeds Assessment of Neuropathic Symptoms and Signs (S-LANSS), which was developed for epidemiological studies [5], and the interview-only Douleur Neuropathique en 4 questions (DN4-interview), which was suggested as being suitable for epidemiological studies [6], both have sensitivities and specificities greater than 0.7, but less than 0.9 in their validation populations. That is, they are imperfect diagnostic tools.

When the prevalence of a disease is low (e.g., 6.9% to 10% of the adult population is estimated to have neuropathic pain) [4], and screening tool sensitivity and specificity are moderate to high (e.g., tools developed to screen for pain with neuropathic features), then the number of false positives is typically numerically greater than the number of false negatives, and prevalence estimates are inflated. From a frequentist statistical standpoint, prevalence estimates can be corrected for the use of imperfect screening tools using the Rogan-Gladen estimator [7]. However, this estimator has significant limitations, such as it assumes fixed sensitivity and specificity values for a questionnaire across populations, and at prevalence extremes, it yields invalid results (prevalence estimates less than zero or greater than one) [8,9]. An alternative approach is to use Bayesian estimation, which allows the modelling of the sensitivity and specificity, and the prevalence estimate within the boundaries of zero and one, and does not assume fixed sensitivity and specificity values across different populations. The method also allows the incorporation of prior knowledge on the prevalence in the estimate of true prevalence [8, 9].

Consequently, we conducted a systematic review of the literature on the prevalence of pain with neuropathic features in the general adult population (using regional or country-level studies only), and then applied Bayesian estimation methods to estimate the corrected prevalence of pain with neuropathic features in the general adult population. As a sensitivity analysis, we calculated our prevalence estimates under four prior distributions (one non-informative prior, and three informative priors).

## Methods

The protocol for this review was registered on the PROSPERO database (CRD42023416845), and the results are reported according to the recommendations of the Preferred Reporting Items for Systematic Review and Meta-Analysis (PRISMA) guideline [10]; the checklist is available in the Supplementary file: Section 1.

### Eligibility criteria

Using the Population, Phenomena of Interest and Context (PICo) framework, our research question was, “*What is the prevalence of neuropathic pain in the general adult population?*”, and our selection criteria were as follows:

#### Inclusion criteria

1. An epidemiological study investigating disease (neuropathic pain) prevalence.
2. The study was conducted at the level of a geographical region (e.g., Great Britain), country, or state/province.
3. The study was conducted in the general adult population (≥18 years of age).
4. Neuropathic pain was assessed as a primary or secondary outcome of the study.
5. Neuropathic pain was screened for using the full Douleur Neuropathique en 4 questions (DN4) or the shortened interview only version (DN4-interview) [6]; the Leeds Assessment of Neuropathic Symptoms and Signs (LANSS) [11] or the self-report version of the LANSS (S-LANSS) [5]; or the painDETECT [12] screening tools.
6. The study was published in English.

#### Exclusion criteria

1. The study focused on one type/cause of neuropathic pain only (e.g., painful diabetic neuropathy only).
2. The study was conducted on a sub-population (e.g., presence of neuropathic pain in adults with chronic pain).

### Information sources and search strategy

We queried three electronic databases: PubMed, Scopus, and Web of Science (search strategies in the Supplementary file: Section 2). The initial search was conducted on 14 April 2023 and spanned the period: 1 January 2001 (the year the first screening tool, LANSS, was published) to 31 March 2023. A second search was conducted on 5 February 2025 and spanned the period: 1 April 2023 to 4 February 2025. Reference lists of papers that passed the initial author-abstract screening process (see below) were assessed to identify additional articles. Grey literature sources were not assessed.

### Selection process and data collection

The results returned from each database were uploaded to Covidence (https://www.covidence.org), an online platform for managing systematic reviews, and duplicates were removed using the built-in automatic duplicate removal tool that uses title, year, volume number, and author similarities to detect duplicates. Once duplicate articles had been removed, articles underwent title and abstract screening, followed by article retrieval and full text screening. Finally, the included articles underwent data extraction and quality assessment. The screening processes were performed in duplicate by four reviewers working independently of each other (PRK, TH, NM, PCC). When there were conflicts, a third reviewer from the group made the final decision. Data extraction and quality assessment were also performed in duplicate by two independent reviewers (TH, NM), with PRK checking for consensus and resolving conflicts between the two reviewers.

A copy of the extraction template used in Covidence is available in the Supplementary file: Section 3. Key items extracted were: publication date; study location (country and region/state/province); sample frame; the criteria used to define whether pain was chronic; the neuropathic pain screening tool used and the threshold used for defining caseness; the number of people approached to take part; the number of people recruited and who completed the questionnaires; the number of people with no pain, acute pain, chronic pain, and chronic pain with neuropathic features. Where sample weights were used by the authors, the raw and weighted sample sizes were recorded.

Study quality and risk of bias were assessed on a Covidence template using the JBI Checklist for Prevalence Studies [13, 14]. A copy of the quality assessment template is available in the Supplementary file: Section 4. The tool includes nine questions, each answered “yes”, “no”, or “unclear”. The checklist does not produce a score, and hence, there is no cut-off score for deciding whether a study should be included in a review. Rather, the evidence is weighed over the questions, and a subjective decision to include a paper is made based on the balance of the evidence.

### Effect measure

The effect measure of interest was the prevalence of pain with neuropathic features in the study sample.

### Reporting bias and certainty assessments

No assessment of reporting bias and certainty was made.

### Data analysis

All analyses and plotting were performed in the R statistical environment (v4.6.0) using functions from the following packages: coda [15], dplyr [16], epiR [17], ggplot2 [18], ggthemes [19], prevalence [20], purrr [21], and tidyr [22]. All analysis scripts and the data used in these scripts are available at Zenodo.org: https://zenodo.org/records/23695163?preview=1&token=eyJhbGciOiJIUzUxMiJ9.eyJpZCI6ImUzNDRjMzRiLWJmYTItNDMxYS04NTc1LTE4MDczMTkzNmIwNyIsImRhdGEiOnt9LCJyYW5kb20iOiI4NDU5ZjhiN2JlYzk2OGYyNTc2YmNkN2YzYWE1ZmY4ZCJ9.q66R1q0gHTh49UCU2weOLUaLE0dIfunfOUqfjazOmKcV4JG30ToexQBLqhePtfIooJ92CcUmM706mlHXc-uRUg

The analysis was conducted as follows:

### Apparent prevalence

1. Apparent prevalence (i.e., no correction for screening tool sensitivity and specificity) was calculated using the number of people detected as having pain with neuropathic features (as detected using screening tools) and the total number of people reported to have completed the questionnaires. For the two studies where population weights were applied to the data, the scaled sample sizes were used instead of the raw numbers [23, 24]. Following data extraction, the proportion (with 95% confidence interval; calculated using the exact binomial method) of people with pain with neuropathic features was calculated for each study.

### Corrected prevalence

1. Because the sensitivity and specificity values of a screening tool are not universal, and apply only to the sample of people and context in which the tool was developed [25–28], we generated distributions of possible sensitivity and specificity values for each questionnaire for inclusion in the Bayesian modelling of corrected prevalence (see below). The distributions of possible sensitivity and specificity values for each screening tool were modelled using a beta distribution, with the two shape parameters for each distribution being determined using the “beta.buster” function in the R “epiR” package. In brief, for each published validation of the screening tools used in the studies included in this review, we extracted the point estimates for the sensitivity and specificity of the screening tool at the scoring threshold used in the reviewed studies. These values were specified as the most likely sensitivity and specificity values for a particular screening tool, with 90% confidence that the sensitivity/specificity value was greater than the sensitivity/specificity value - 0.1. For example, the published sensitivity for a score ≥ 19 on the painDETECT instrument is 0.84 [12], which was specified as the most likely value, and we were 90% confident that the sensitivity value was greater than 0.74.
2. We used the “truePrev” function from the R “prevalence” package to calculate the corrected prevalence for each of the reviewed studies. The values specified for the number of people with pain with neuropathic features and the total sample size were the same as those used in the calculation of the apparent prevalences (see above). In addition, the distributions of sensitivity and specificity values for each screening tool were specified using the beta distributions that had been calculated (see above). The data were then modelled under four conditions:

a. A non-informative prior distribution, specified as Beta(1, 1);
b. The informative prior distributions were generated using the “beta.buster” function in the R “epiR” package, with distribution assumptions based on the results of the apparent prevalence point estimates from the systematic review component of the study. The three prior distributions were:

i. Beta(5.619, 42.5712), models a distribution with the most likely prevalence being 10%, with 95% confidence that the true prevalence is less than 20%;
ii. Beta(2.063, 21.197), models a distribution with the most likely prevalence being 5%, with 95% confidence that the true prevalence is less than 20%;
iii. Beta(1.398, 16.522), models a distribution with the most likely prevalence being 2.5%, with 95% confidence that the true prevalence is less than 20%.

Plots of the three informative prior distributions are provided in Section 5 of the Supplementary file.

The modelling started with 5 chains using a burn-in of 10,000 iterations, and an update of 100,000 iterations. The convergence of each model was then checked using the Brooks-Gelman-Rubin statistic, with convergence accepted at values < 1.2. If convergence was not attained, the burn-in and update iterations were each increased by a factor of 10 until convergence was achieved. The results of the Bayesian estimates of the corrected prevalence for each study were then plotted as the point estimate (median) with 95% high-density intervals.

## Results

### Search results

Figure 1 provides an overview of the article selection process. After duplicate removal and title-abstract screening, 21 articles underwent full-text screening. Twelve of these 21 studies were deemed not to fit the inclusion criteria; a list of the 12 studies and the reason for their exclusion is provided in Section 6 of the Supplementary file [2, 3, 29–38]. The remaining 9 studies underwent data extraction [23, 24, 39–45]. However, during the extraction process, it became evident that sensitivity and specificity data for the screening tool used in one of the studies (Japanese version of the painDETECT) were not available [45]. Only the specificity of the tool was forthcoming after emailing the authors, so the study was excluded at the data extraction phase.

**Figure 1.**
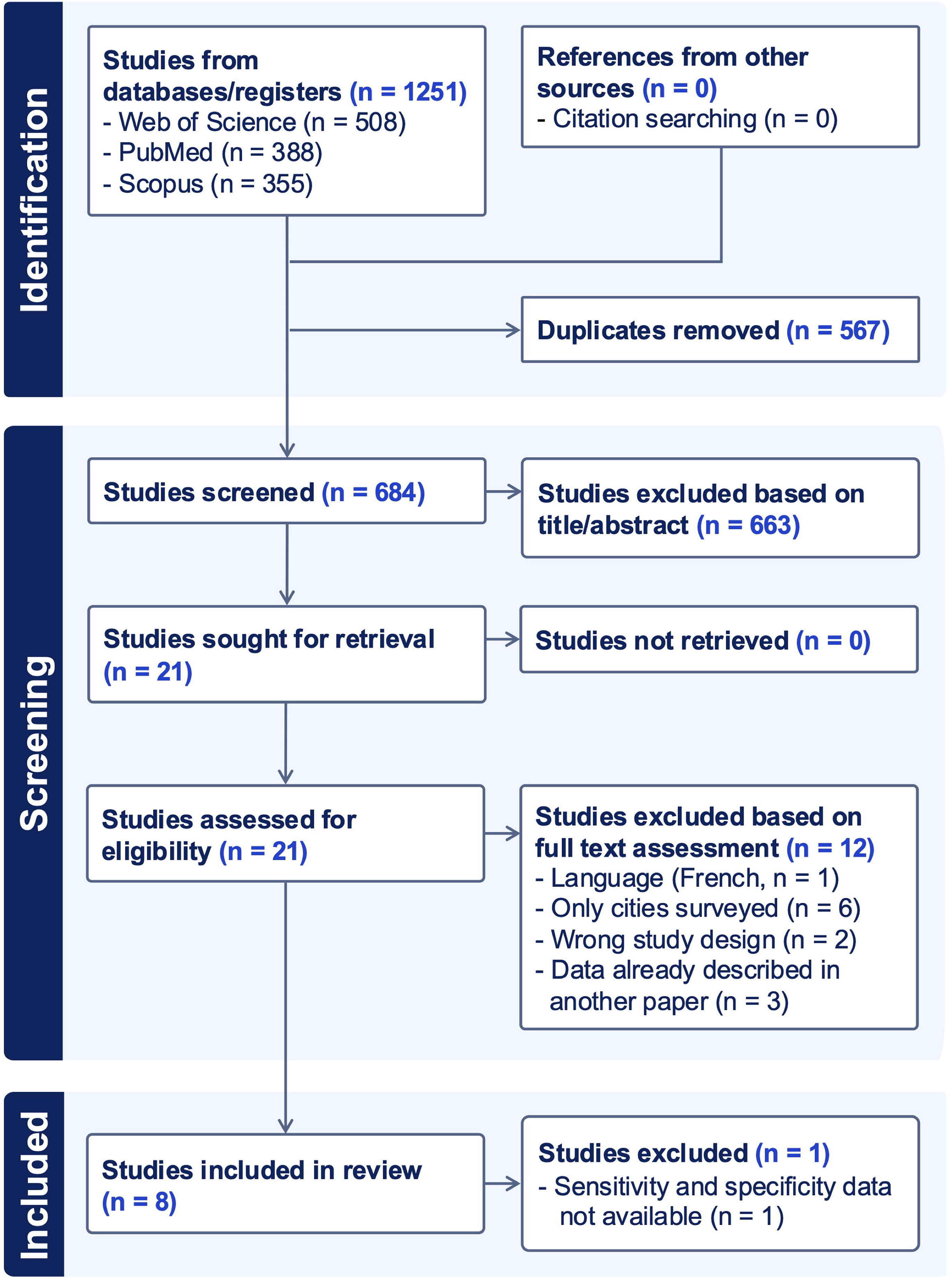
PRISMA flow diagram for the study selection process

### Study characteristics

The characteristics of the eight studies that were included in the review are provided in Table 1. One of the studies, VanDenKerkhof et al., 2016 [24], screened the recruited population for the presence of chronic pain with neuropathic features using the DN4-interview and the S-LANSS; we decided to treat the results from the two screening tools as two independent datasets, with both datasets being included in our analyses.

**Table 1.** Description of the studies included in the systematic review and meta-analysis (eight studies, with one study having two datasets)

| **Article** | **Country (region)** | **Screening tool** | **Sensitivity of the screening tool at the threshold score** | **Specificity of the screening tool at the threshold score** | **Screening tool threshold score for defining the presence of pain with neuropathic features** | **Additional requirements for the case definition of chronic pain with neuropathic features** | **Number of individuals contacted** | **Number of individuals recruited<br>[response rate]<sup>h</sup> (%)** | **Analytical set: Number of individuals completing the screening tool assessment (scaled N)<br>[completion rate]<sup>i</sup> (%)** | **Number of individuals in the analytical set with chronic pain with neuropathic features (scaled N)** |
|---|---|---|---|---|---|---|---|---|---|---|
| Baskozos et al., 2023 | UK (regions: England, Scotland, Wales) | DN4 (interview)<sup>e</sup> | 0.78 | 0.812 | ≥ 3 | Pain duration ≥ 3 months | 335,587 | 167,203<br>[49.8] | 148,828<br>[44.3] | 13,744 |
| Bouhassira et al., 2008<sup>a</sup> | France (national) | DN4 (interview)<sup>e</sup> | 0.78 | 0.812 | ≥ 3 | Pain duration ≥ 3 months | 30,155 | 24,497<br>[81.2] | 23,712<br>[78.6] | 1,631 |
| DiBonaventura et al., 2017 | USA (national) | PainDETECT | 0.84 | 0.84 | ≥ 19 | Pain duration ≥ 3 months | ~1,000,000<sup>g</sup> | 24,925<br>[2.5] | 24,925<br>(241,829,594)<br>[2.5] | 2,548<br>(24,206,788) |
| Harifi et al., 2013<sup>b</sup> | Morocco (national) | DN4 (interview)<sup>e</sup> [Arabic version] | 0.894 | 0.724 | ≥ 3 | Pain duration ≥ 3 months | 8,382 | 5,328<br>[63.6] | 5,116<br>[61.0] | 542 |
| Hebert et al., 2021<sup>c</sup> | UK (region: Scotland) | DN4 (interview)<sup>e</sup> | 0.78 | 0.812 | ≥ 3 | Pain duration ≥ 3 months | 20,221 | 7,240<br>[35.8] | 6,058<br>[30.0] | 932 |
| Toth et al., 2009 | Canada (region: Alberta) | DN4 (interview)<sup>e</sup> | 0.78 | 0.812 | ≥ 3 | Pain duration ≥ 3 months | 4,167 | 1,207<br>[29.0] | 1,207<br>[29.0] | 216 |
| VanDenKerkhof et al., 2016<sup>d</sup><br>Dataset 1 | Canada (national) | DN4 (interview)<sup>e</sup> | 0.78 | 0.812 | ≥ 3 | Pain duration ≥ 3 months<br>+ likely or possible cause of neuropathic pain | 7,134 | 1,504<br>[21.1] | 1,504<br>(26,423,076)<br>[21.1] | 166<br>(3,027,521) |
| VanDenKerkhof et al., 2016<sup>d</sup><br>Dataset 2 | Canada (national) | S-LANSS (self-complete)<sup>f</sup> | 0.74 | 0.76 | ≥ 12 | Pain duration ≥ 3 months<br>+ likely or possible cause of neuropathic pain | 7,134 | 1,504<br>[21.1] | 1,504<br>(26,423,076)<br>[21.1] | 123<br>(2,030,351) |
| Yawn et al., 2009 | USA (region: Olmsted County, Minnesota) | S-LANSS (self-complete)<sup>f</sup> | 0.74 | 0.76 | ≥ 12 | Pain duration ≥ 3 months | 5,897 | 3,575<br>[60.6] | 3,575<br>[60.6] | 315 |

a. Population weights were used for the analysis of demographic data, but their use for pain prevalence calculations was unclear, so raw data were used to calculate prevalence
b. The number of cases back-calculated from the reported prevalence
c. The number of individuals in the analytical set and the number of individuals with pain with neuropathic features were extracted from Table 7
d. Raw numbers were extracted from Figure 1. Weighted numbers were extracted from Table 1
e. DN4: Douleur Neuropathique en 4 Questions (interview only)
f. S-LANSS: Self-completed Leeds Assessment of Neuropathic Symptoms and Signs
g. The exact number was not reported
h. Response rate: Number of individuals recruited / number of individuals approached to take part in the study
i. Completion rate: Number of respondents included in the analysis/number of individuals approached to take part in the study

One study each was conducted in France [44] and in Morocco [40], and two studies were conducted in each of Canada [24, 42], the United Kingdom [39, 41], and the United States of America [23, 43]. Five studies were national studies [23, 24, 39, 40, 44], with the rest being regional. Six studies used the DN4-interview [24, 39–42, 44], two studies used the S-LANSS [24, 43], and one study used the painDETECT [23]. In all cases, the published recommended screening tool scores were used to define whether a participant had pain with neuropathic features (DN4-interview: ≥ 3 [6, 46], painDETECT: ≥ 19 [12], and S-LANSS: ≥ 12 [5]). Similarly, all studies used pain lasting ≥ 3 months to classify whether pain was chronic. The study by VanDenKerkhof et al. [24] included the additional criterion that the pain was associated with a likely (e.g., accident with nerve damage, amputation, and shingles) or probable (e.g., surgery more than 3 months ago, back problem, diabetes, cancer) insult to the somatosensory nervous system that could result in neuropathic pain. The point estimates of the sensitivity and specificity of the three screening tools were extracted from their respective validation studies (i.e., DN4-interview in French [6], DN4-interview in Arabic [46], S-LANSS in English [5]). Where no validation study was available for the language version of the screening tool used in a survey, the sensitivity and specificity values were taken from the original validation study (e.g., an unvalidated English translation of the original French version of the DN4-interview was used in four of the surveys [24, 39, 41, 42], so the sensitivity and specificity reported in the French validation study were used [6]).

In total, across the 8 studies, approximately 1,411,552 people were approached to participate in the surveys (largest: ∼1,000,000 [23], smallest: 4,167 [42]). Of those approached, 235,479 people responded (average response rate: 16.7%; lowest: 2.5% [23], highest: 81.2% [44]), and 214,925 people had complete data that could be analyzed (average completion rate: 15.2%; lowest: 2.5% [23], highest: 78.6% [44]). Across the 9 datasets (VanDenKerkhof et al. [24] contributed 2 datasets), the total number of people fitting the criteria for chronic pain with neuropathic features was 20,094 when using was VanDenKerkhof’s DN4-interview results, and 20,051 when using VanDenKerkhof’s S-LANSS results. After rounding, the raw prevalence for both totals was 9.3% (95% confidence interval: 9.2 to 9.5).

### Study quality and risk of bias

Results of the JBI Checklist for Prevalence Studies are provided in Table 2. Overall, there were limitations in several quality metrics, particularly the reporting of standardization of methods for questionnaire administration, the appropriateness of the statistical analysis, and low response rates. No studies were excluded based on the overall balance of the evidence quality. However, based on the low number of studies and the short-comings of some of the studies, we did not perform a meta-analysis on the data.

**Table 2.**
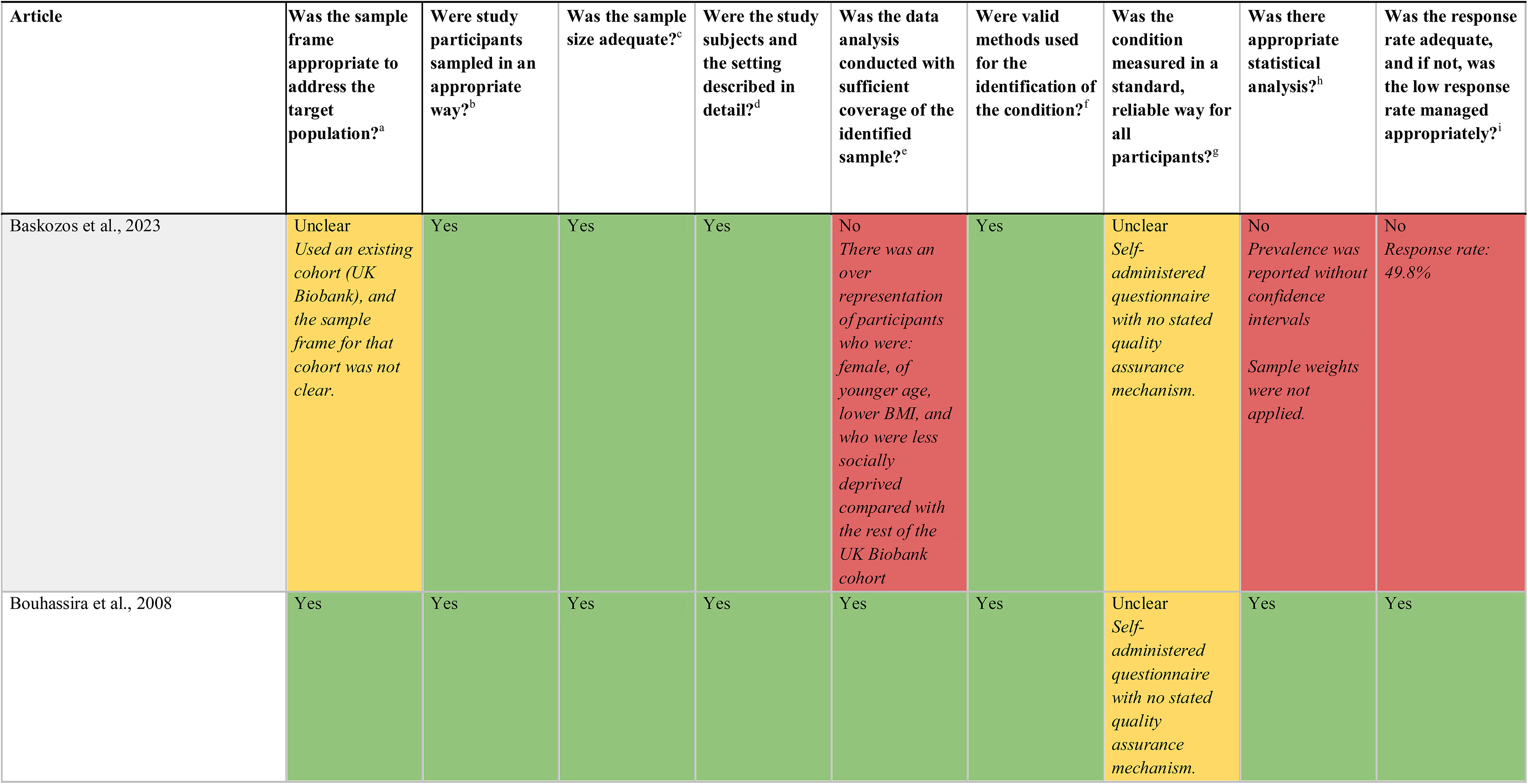
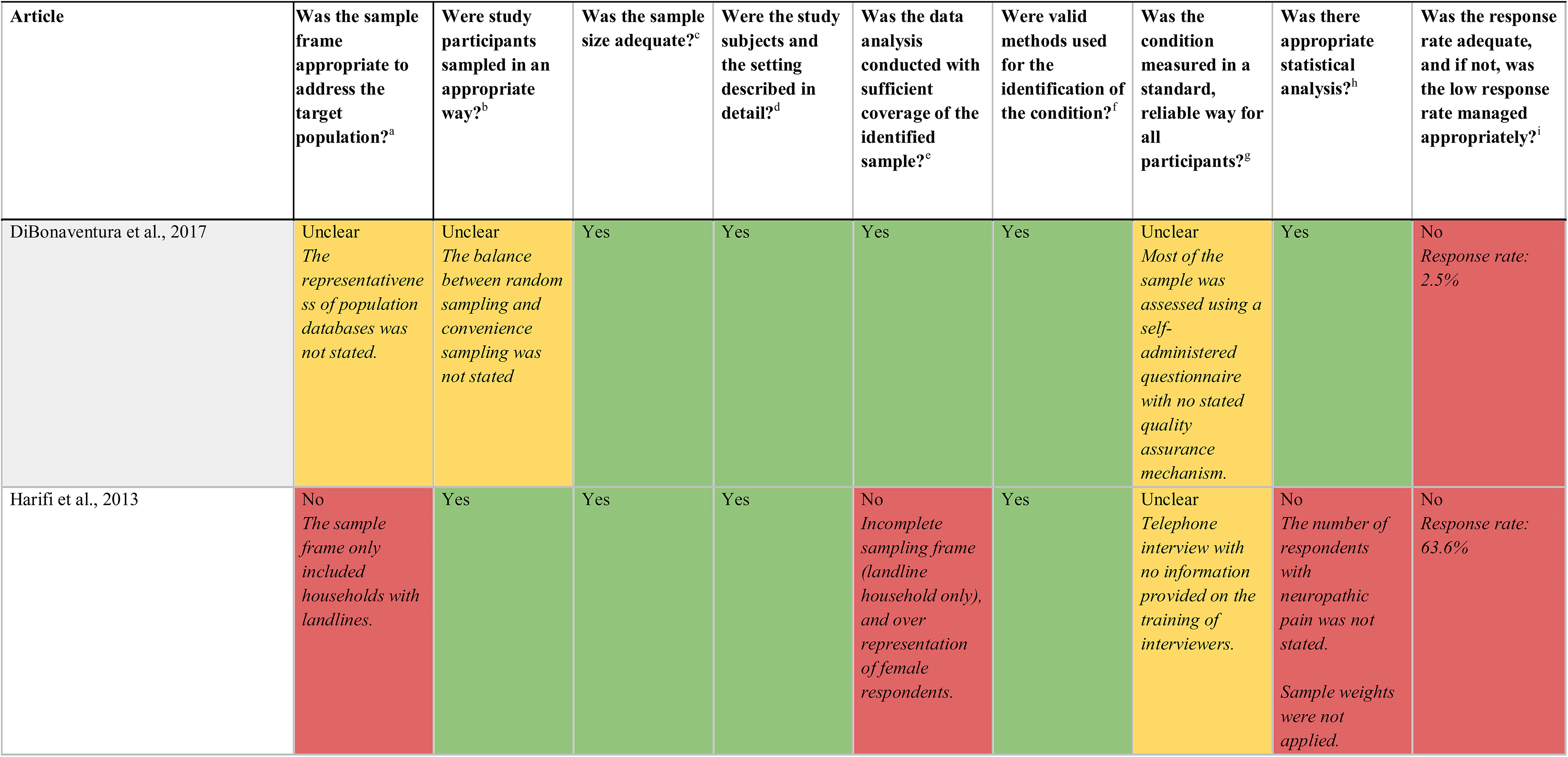
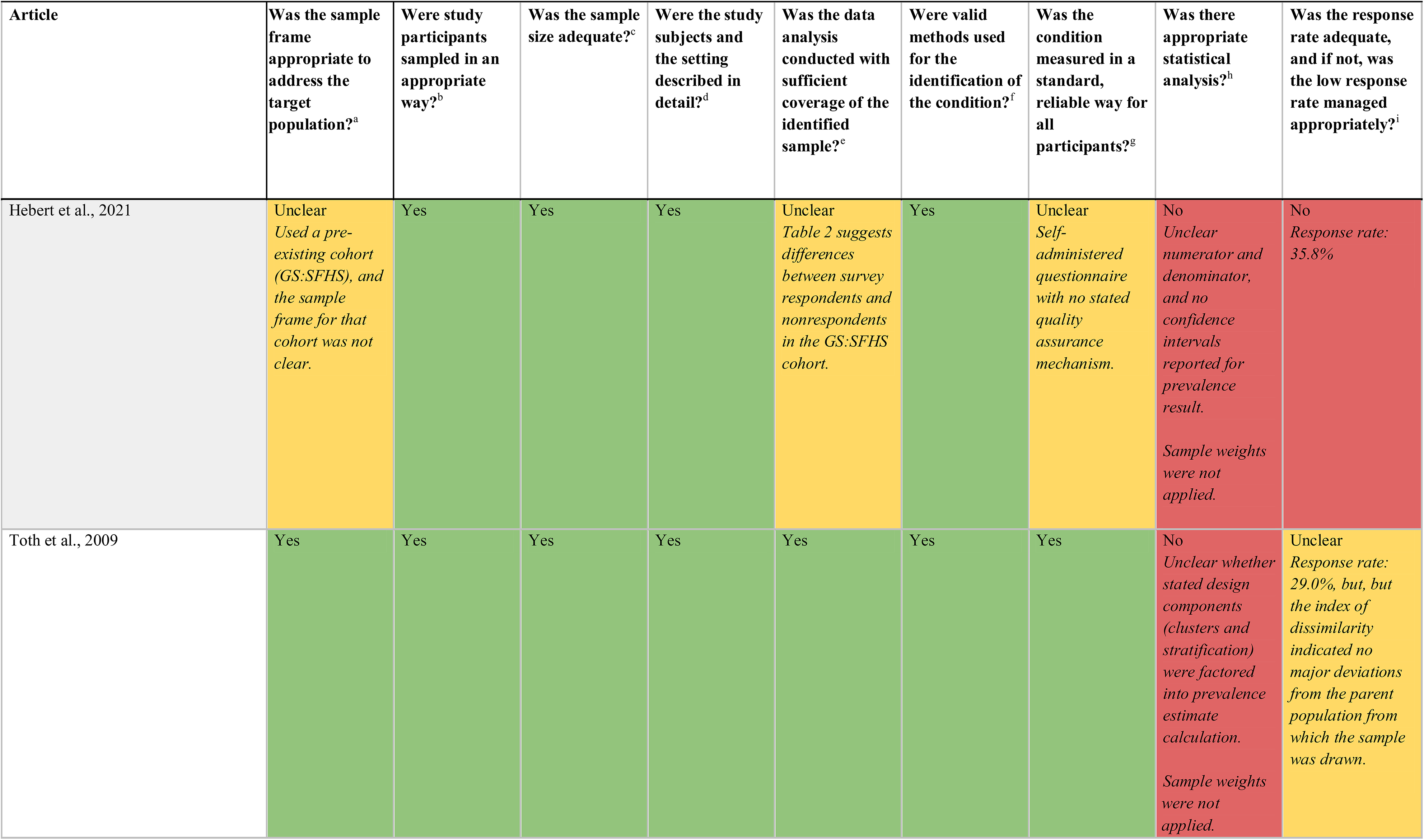
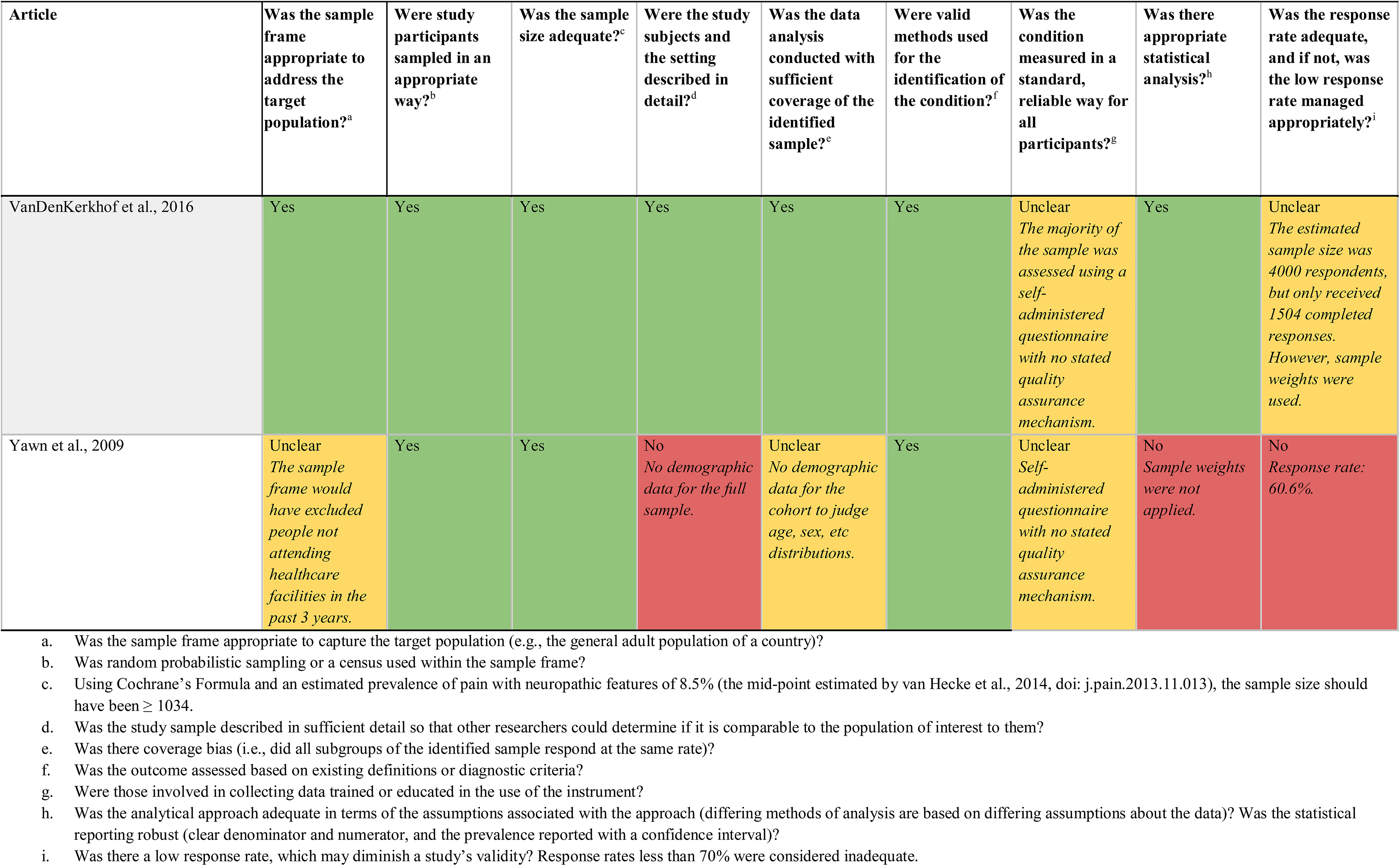
Quality assessment of the studies included in the systematic review and meta-analysis (eight studies)

| Article | Was the sample frame appropriate to address the target population?<sup>a</sup> | Were study participants sampled in an appropriate way?<sup>b</sup> | Was the sample size adequate?<sup>c</sup> | Were the study subjects and the setting described in detail?<sup>d</sup> | Was the data analysis conducted with sufficient coverage of the identified sample?<sup>e</sup> | Were valid methods used for the identification of the condition?<sup>f</sup> | Was the condition measured in a standard, reliable way for all participants?<sup>g</sup> | Was there appropriate statistical analysis?<sup>h</sup> | Was the response rate adequate, and if not, was the low response rate managed appropriately?<sup>i</sup> |
|---|---|---|---|---|---|---|---|---|---|
| Baskozos et al., 2023 | Unclear<br>*Used an existing cohort (UK Biobank), and the sample frame for that cohort was not clear.* | Yes | Yes | Yes | No<br>*There was an over representation of participants who were: female, of younger age, lower BMI, and who were less socially deprived compared with the rest of the UK Biobank cohort* | Yes | Unclear<br>*Self-administered questionnaire with no stated quality assurance mechanism.* | No<br>*Prevalence was reported without confidence intervals*<br>*Sample weights were not applied.* | No<br>*Response rate: 49.8%* |
| Bouhassira et al., 2008 | Yes | Yes | Yes | Yes | Yes | Yes | Unclear<br>*Self-administered questionnaire with no stated quality assurance mechanism.* | Yes | Yes |
| Article | Was the sample frame appropriate to address the target population?<sup>a</sup> | Were study participants sampled in an appropriate way?<sup>b</sup> | Was the sample size adequate?<sup>c</sup> | Were the study subjects and the setting described in detail?<sup>d</sup> | Was the data analysis conducted with sufficient coverage of the identified sample?<sup>e</sup> | Were valid methods used for the identification of the condition?<sup>f</sup> | Was the condition measured in a standard, reliable way for all participants?<sup>g</sup> | Was there appropriate statistical analysis?<sup>h</sup> | Was the response rate adequate, and if not, was the low response rate managed appropriately?<sup>i</sup> |
| DiBonaventura et al., 2017 | Unclear<br>*The representativeness of population databases was not stated.* | Unclear<br>*The balance between random sampling and convenience sampling was not stated* | Yes | Yes | Yes | Yes | Unclear<br>*Most of the sample was assessed using a self-administered questionnaire with no stated quality assurance mechanism.* | Yes | No<br>*Response rate: 2.5%* |
| Harifi et al., 2013 | No<br>*The sample frame only included households with landlines.* | Yes | Yes | Yes | No<br>*Incomplete sampling frame (landline household only), and over representation of female respondents.* | Yes | Unclear<br>*Telephone interview with no information provided on the training of interviewers.* | No<br>*The number of respondents with neuropathic pain was not stated.*<br>*Sample weights were not applied.* | No<br>*Response rate: 63.6%* |
| Article | Was the sample frame appropriate to address the target population?<sup>a</sup> | Were study participants sampled in an appropriate way?<sup>b</sup> | Was the sample size adequate?<sup>c</sup> | Were the study subjects and the setting described in detail?<sup>d</sup> | Was the data analysis conducted with sufficient coverage of the identified sample?<sup>e</sup> | Were valid methods used for the identification of the condition?<sup>f</sup> | Was the condition measured in a standard, reliable way for all participants?<sup>g</sup> | Was there appropriate statistical analysis?<sup>h</sup> | Was the response rate adequate, and if not, was the low response rate managed appropriately?<sup>i</sup> |
| Hebert et al., 2021 | Unclear<br>*Used a pre-existing cohort (GS:SFHS), and the sample frame for that cohort was not clear.* | Yes | Yes | Yes | Unclear<br>*Table 2 suggests differences between survey respondents and nonrespondents in the GS:SFHS cohort.* | Yes | Unclear<br>*Self-administered questionnaire with no stated quality assurance mechanism.* | No<br>*Unclear numerator and denominator, and no confidence intervals reported for prevalence result.*<br>*Sample weights were not applied.* | No<br>*Response rate: 35.8%* |
| Toth et al., 2009 | Yes | Yes | Yes | Yes | Yes | Yes | Yes | No<br>*Unclear whether stated design components (clusters and stratification) were factored into prevalence estimate calculation.*<br>*Sample weights were not applied.* | Unclear<br>*Response rate: 29.0%, but, but the index of dissimilarity indicated no major deviations from the parent population from which the sample was drawn.* |
| Article | Was the sample frame appropriate to address the target population?<sup>a</sup> | Were study participants sampled in an appropriate way?<sup>b</sup> | Was the sample size adequate?<sup>c</sup> | Were the study subjects and the setting described in detail?<sup>d</sup> | Was the data analysis conducted with sufficient coverage of the identified sample?<sup>e</sup> | Were valid methods used for the identification of the condition?<sup>f</sup> | Was the condition measured in a standard, reliable way for all participants?<sup>g</sup> | Was there appropriate statistical analysis?<sup>h</sup> | Was the response rate adequate, and if not, was the low response rate managed appropriately?<sup>i</sup> |
| VanDenKerkhof et al., 2016 | Yes | Yes | Yes | Yes | Yes | Yes | Unclear<br>*The majority of the sample was assessed using a self-administered questionnaire with no stated quality assurance mechanism.* | Yes | Unclear<br>*The estimated sample size was 4000 respondents, but only received 1504 completed responses. However, sample weights were used.* |
| Yawn et al., 2009 | Unclear<br>*The sample frame would have excluded people not attending healthcare facilities in the past 3 years.* | Yes | Yes | No<br>*No demographic data for the full sample.* | Unclear<br>*No demographic data for the cohort to judge age, sex, etc distributions.* | Yes | Unclear<br>*Self-administered questionnaire with no stated quality assurance mechanism.* | No<br>*Sample weights were not applied.* | No<br>*Response rate: 60.6%.* |

a. Was the sample frame appropriate to capture the target population (e.g., the general adult population of a country)?
b. Was random probabilistic sampling or a census used within the sample frame?
c. Using Cochrane’s Formula and an estimated prevalence of pain with neuropathic features of 8.5% (the mid-point estimated by van Hecke et al., 2014, doi: j.pain.2013.11.013), the sample size should have been ≥ 1034.
d. Was the study sample described in sufficient detail so that other researchers could determine if it is comparable to the population of interest to them?
e. Was there coverage bias (i.e., did all subgroups of the identified sample respond at the same rate)?
f. Was the outcome assessed based on existing definitions or diagnostic criteria?
g. Were those involved in collecting data trained or educated in the use of the instrument?
h. Was the analytical approach adequate in terms of the assumptions associated with the approach (differing methods of analysis are based on differing assumptions about the data)? Was the statistical reporting robust (clear denominator and numerator, and the prevalence reported with a confidence interval)?
i. Was there a low response rate, which may diminish a study’s validity? Response rates less than 70% were considered inadequate.

### Modelling results

The results are reported according to the screening tool used [DN4: Figure 2 and Table S1 (Supplementary file: Section 7); S-LANSS: Figure 3 and Table S2 (Supplementary file: Section 7); painDETECT: Figure 4 and Table S3 (Supplementary file: Section 7)].s

**Figure 2.**
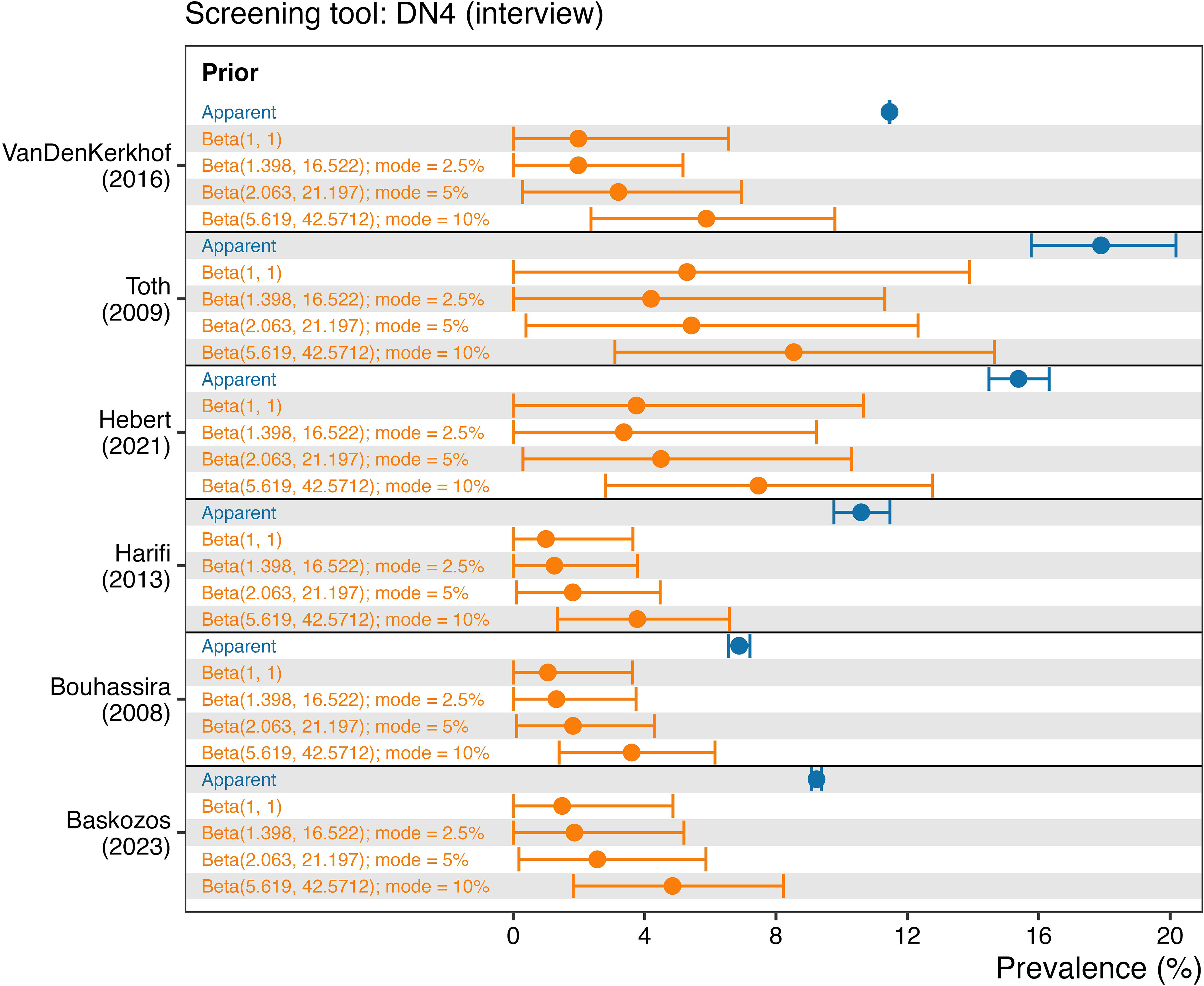
Apparent (blue) and posterior distributions (orange) of the prevalence of pain with neuropathic characteristics in six studies using the Douleur Neuropathique en 4 - interview (DN4-interview) questionnaire. For each study, apparent prevalence is shown as the point estimate with 95% confidence intervals, while posterior distributions are shown as 95% high-density intervals with point estimates. Posterior distributions were generated under four different priors [a noninformative prior: Beta(1, 1), an informative prior with a mode of 2.5%: Beta(1.398, 16.522), an informative prior with a mode of 5%: Beta(2.063, 21.197), and an informative prior with a mode of 10%: Beta(5.619, 42.5712)]

**Figure 3.**
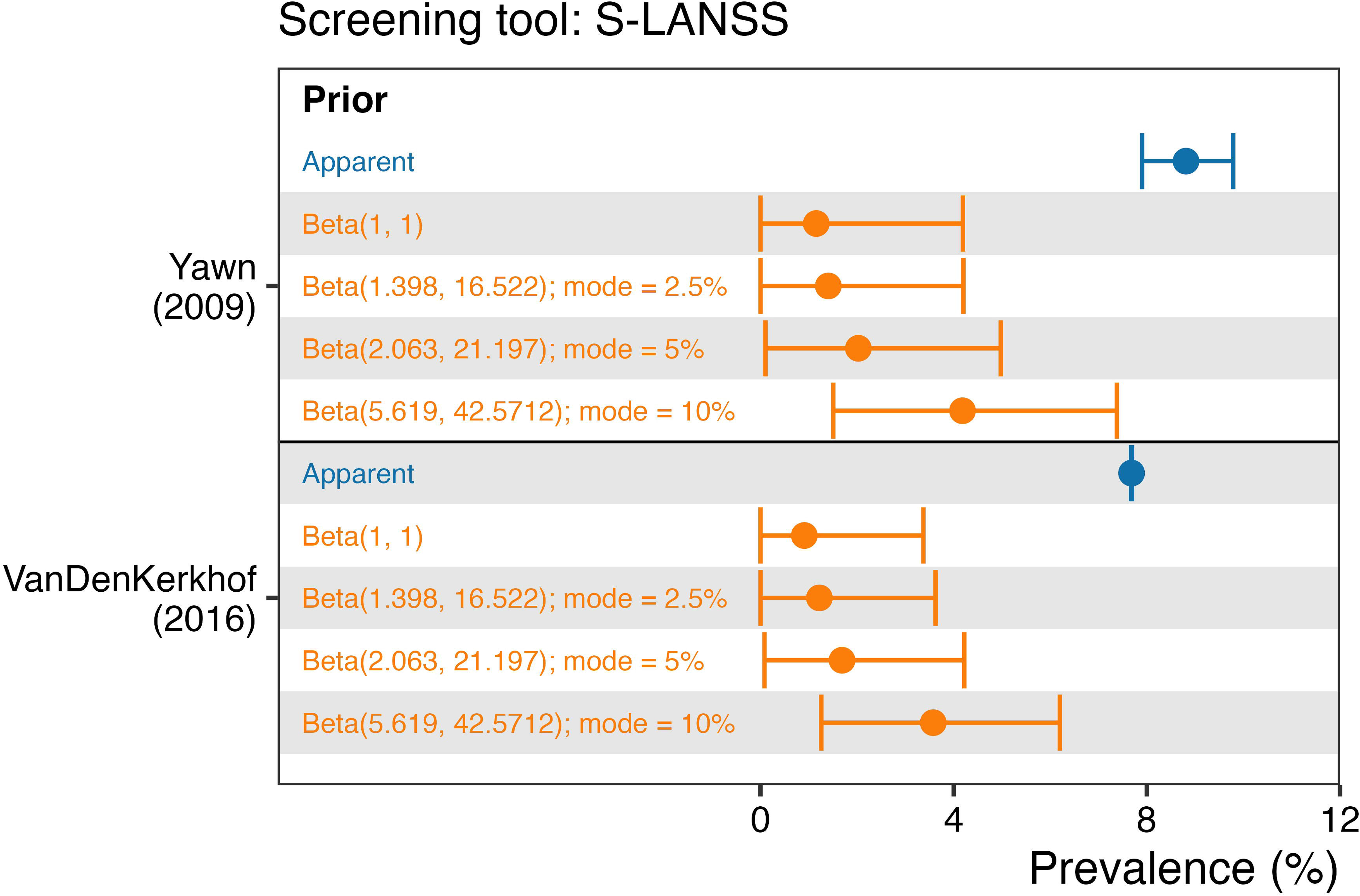
Apparent (blue) and posterior distributions (orange) of the prevalence of pain with neuropathic characteristics in two studies using the Self-complete Leeds Assessment of Neuropathic Symptoms and Signs (S-LANSS) questionnaire. For each study, apparent prevalence is shown as the point estimate with 95% confidence intervals, while posterior distributions are shown as 95% high-density intervals with point estimates. Posterior distributions were generated under four different priors [a noninformative prior: Beta(1, 1), an informative prior with a mode of 2.5%: Beta(1.398, 16.522), an informative prior with a mode of 5%: Beta(2.063, 21.197), and an informative prior with a mode of 10%: Beta(5.619, 42.5712)]

**Figure 4.**
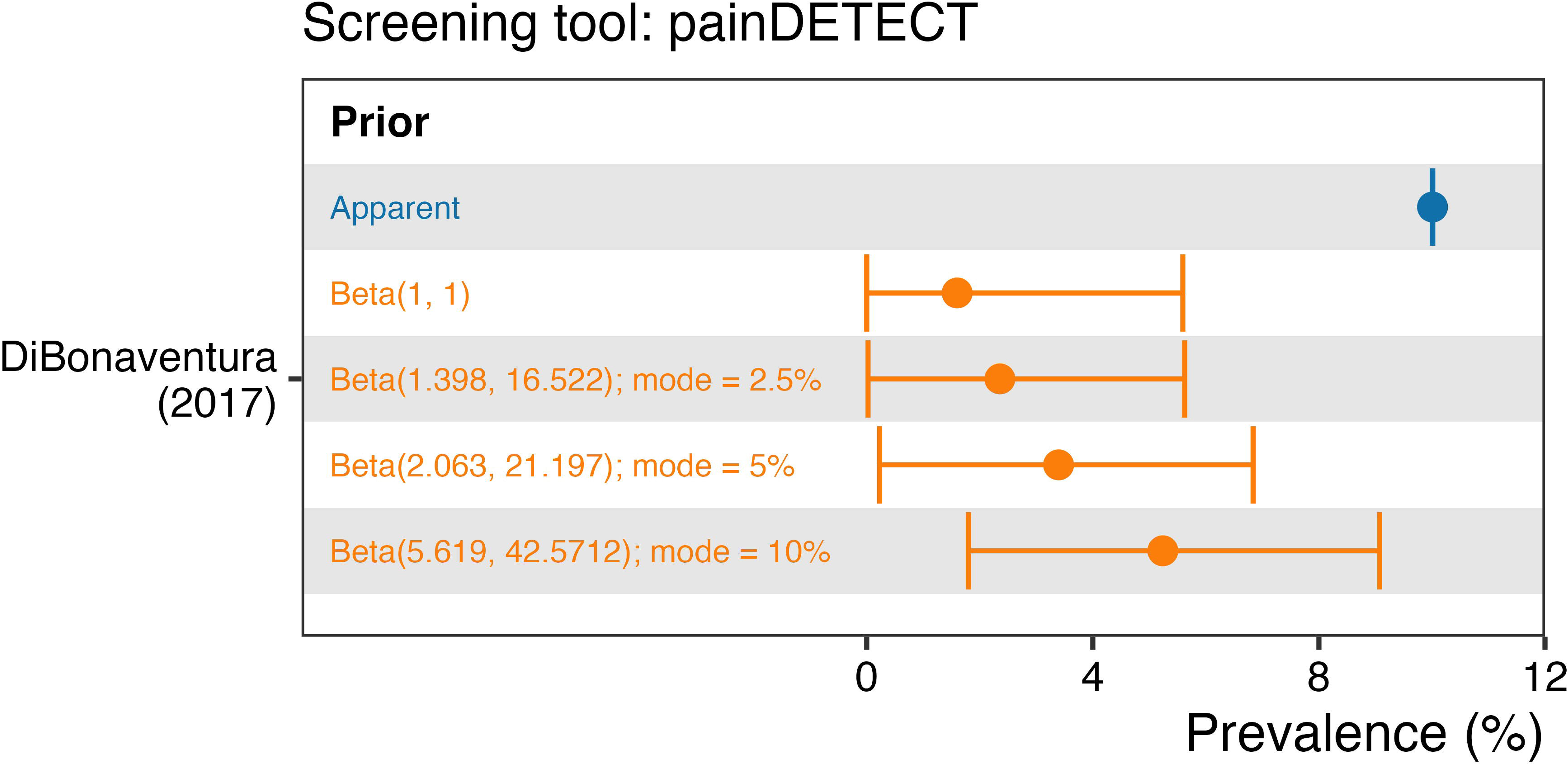
Apparent (blue) and posterior distributions (orange) of the prevalence of pain with neuropathic characteristics in the single study using the painDETECT questionnaire. Apparent prevalence is shown as the point estimate with 95% confidence intervals, while posterior distributions are shown as 95% high-density intervals with point estimates. Posterior distributions were generated under four different priors [a noninformative prior: Beta(1, 1), an informative prior with a mode of 2.5%: Beta(1.398, 16.522), an informative prior with a mode of 5%: Beta(2.063, 21.197), and an informative prior with a mode of 10%: Beta(5.619, 42.5712)]

Overall, precision of the posterior prevalence estimates was fair except for the studies by Toth ([42], DN4-interview, Canada) and by Hébert ([41], DN4-interview, Scotland), where there was a high level of imprecision (wide 95% high-density intervals; > 10 percentage points).

Importantly, within each of the eight studies (nine datasets), there was high consistency in the point estimates (medians) of the posterior distributions, and considerable overlap in the 95% high-density intervals, for the non-informative prior and the informative priors with prevalences centered at 2.5% and 5%. The median spread in posterior point estimates across these three priors, for the nine datasets, was 1.1 percentage points, with the narrowest spread being 0.7 percentage points (from 1.1% to 1.8%; Bouhassira [44], DN4-interview, France), and the widest spread being 1.8 percentage points (from 1.6% to 3.4%; DiBonaventura [23], painDETECT, USA). The median (range) of the prevalence estimates for the non-informative prior was 1.5% (0.9 to 5.3), for the informative prior centered at 2.5% prevalence it was 1.9% (1.2 to 4.2), and for the informative prior centered at 5% prevalence it was 2.6% (1.7 to 5.4). This consistency of results across the first three priors suggests that the posterior inference is generally robust to plausible prior specifications within the lower-prevalence range.

However, posterior estimates obtained using the prior centered on 10% prevalence were shifted towards higher values, indicating that the available data did not completely override this stronger prior assumption; the median (range) of the prevalence estimate was 4.9% (3.6 to 8.5). Nevertheless, irrespective of the questionnaire administered in a study or the prior specified in the Bayesian modelling, all posterior median prevalences were far less than the apparent prevalence estimates. While apparent prevalence point estimates ranged across 11 percentage points from a minimum of 6.9% [44] to a maximum of 17.9% [42], the corrected, Bayesian posterior point estimates ranged across 7.6 percentage points from a minimum of 0.9% [40] to a maximum of 8.5% [42]. In summary, all Bayesian models produced consistent, and substantial, downward revisions of the apparent prevalences reported in the nine datasets. The median (range) of the difference between the apparent point estimate for each study and the corresponding Bayesian posterior estimate for the non-informative prior was −8.4 percentage points (−12.6 to −5.8), for the informative prior centered at 2.5% prevalence it was −7.7 percentage points (−13.7 to −5.6), for the informative prior centered at 5% prevalence it was −6.8 percentage points (−12.5 to −5.1), and for the informative prior centered at 10% prevalence it was −4.9 percentage points (−9.4 to −3.3).

## Discussion

We undertook to provide estimates of the prevalence of pain with neuropathic characteristics in the general adult population by correcting the apparent prevalences retrieved from a systematic review using Bayesian models that account for the use of imperfect disease screening tools. Correcting for screening tool sensitivity and specificity shifted the prevalence estimates substantially lower across all eight studies (nine datasets) retrieved by the systematic review. These downward shifts in prevalence estimates were robust to the prior that was specified such that the non-informative prior, and the informative priors with distributions centered on a disease prevalence of 2.5% and 5% produced very consistent posterior point estimates and high-density intervals. Even for the prior with the distribution centered at a 10% disease prevalence, the shift in prevalence estimate was meaningful, with a median reduction in prevalence estimation of about 5 percentage points.

A potential limitation of our analytical approach was our choice of informative priors. Because we could not be sure of the true prevalence of pain with neuropathic characteristics, we used three informative priors and examined the effect of those priors on the posterior estimates. In all three cases, we assumed that there was a 95% probability that the prevalence of pain with neuropathic characteristics was less than 20%. This assumption was based on the results of Toth et al. [42], who reported the greatest estimate of apparent neuropathic pain prevalence in a general population survey, namely, 17.9% (95% confidence interval:15.8 to 20.2), and the results of the last systematic review on the topic, which estimated an upper range of 10% for the apparent prevalence [4]. This latter study also informed us on the mode for one of the prior probability distributions of 10%. The prior distributions with modes of 5% and 2.5% prevalence then stemmed from this informative prior; halving the mode of the prior at each step. We feel that our informative priors cover the prevalence expected from the apparent prevalence estimates (∼10%), and reductions in prevalence anticipated from the Bayesian modelling (2.5% and 5%). Given the stability of the estimates across the informative (and the non-informative) priors, with only the prior with a mode of 10% prevalence showing some deviation from the others, we believe our choice of informative priors was appropriate.

Our modelling was designed to correct the apparent prevalence of pain with neuropathic characteristics based on the use of imperfect screening tools (i.e., tool sensitivity and specificity < 1), yet we did not treat the published sensitivity and specificity of the screening tools as fixed values in our models. Instead, we modelled the sensitivity and specificity values themselves. Our reasoning for modelling these values was that all three tools were used outside of the context in which they were first validated. For example, the sensitivity and specificity values for each of the tools were generated from samples of healthcare-seeking individuals with pain, not general population samples. Also, the painDETECT was validated for use in people with chronic low back pain only, and the sensitivity and specificity values are for the German-language version [12], not the English version used by DiBonaventura and colleagues [23]. Similarly, the English version of the DN4-interview was used in four of the studies we reviewed [24, 39, 41, 42], but sensitivity and specificity data are not available for the English version (we based our modelling on the French-language validation data). It is known that the validation sample and the context influence the sensitivity and specificity of a screening tool [25–28], and therefore it stands to reason that using fixed values for screening tool sensitivity and specificity is an unreasonable assumption, and that the sensitivity and specificity values should themselves be modelled. In our modelling of screening tool sensitivity and specificity, we made sure not to discard the published values by stipulating that the published values were the most likely sensitivity and specificity values for a particular screening tool, with 90% confidence that the sensitivity/specificity value was greater than the sensitivity/specificity value - 0.1.

While our analysis approach corrects for the use of imperfect screening tools, inherent deficiencies across all the included studies also could have led to positive reporting bias in the original studies (Table 2). That is, there was a high prevalence of unclear or deficient sampling frames, unclear or absent controls for coverage bias and adjustment to reference populations, and response rates were generally low to very low. Moreover, screening tools such as the DN4-interview, S-LANSS, and painDETECT are all heavily reliant on pain symptomology, but while the co-occurrence of certain pain symptoms can be suggestive that a pain is neuropathic in origin, these symptom clusters are not pathognomic. Indeed, in the clinical context (as opposed to the epidemiological context) [47], a combination of particular co-occurring symptoms, a relevant medical history, and a plausible neuroanatomical distribution are all required for the grading of a pain as being possible neuropathic pain; the lowest level of diagnostic certainty [48]. One study in our sample factored causes of neuropathic pain into their case definition [24], but that still means that none of the studies met the minimum level of evidence to define the presence of possible neuropathic pain instead of pain with neuropathic characteristics. We propose that any epidemiological studies to estimate the prevalence of neuropathic pain as such, and not pain with neuropathic features, must include not only symptom screens, but also solicit information on medical history (e.g., the presence of a preceding injury or disease of the somatosensory nervous system that could explain the pain) and site of pain (e.g., body charts) [47].

VanDenKerkhof and colleagues [24] assessed neuropathic pain using two screening tools, the DN4-interview and the S-LANSS, and the study therefore provides interesting insight into differences in estimates when using different screening tools for case ascertainment in the same population. The point estimates for apparent prevalence were 7.7% (S-LANSS) and 11.5% (DN4-interview), and the direction of the difference in estimates was maintained, but to a much lower magnitude, across the suite of Bayesian-corrected point estimates [noninformative prior: 0.9% (S-LANSS) vs 2% (DN4-interview); informative prior with a mode of 2.5%: 1.2% (S-LANSS) vs 2% (DN4-interview); informative prior with a mode of 5%: 1.7% (S-LANSS) vs 3.2% (DN4-interview); and informative prior with a mode of 10%: 3.6% (S-LANSS) vs 5.9% (DN4-interview)]. The question then arises as to which of the screening tools gives the most accurate estimate of disease prevalence? The DN4-interview relies purely on the assessment of the presence/absence of symptoms related to the pain quality and associated sensory symptoms [6], while the S-LANSS assesses pain quality, associated symptoms, changes in skin color, and a self-examination of the painful and a non-painful area using static pressure (pressing with a finger) to assess for numbness, and dynamic touch (gentle rubbing of the skin) to assess for evoked symptoms [5]. While the sensitivity and specificity of the S-LANSS (sensitivity: 0.74, specificity: 0.76) is less than those of the DN4-interview (sensitivity: 0.78, specificity: 0.81), the S-LANSS’s validation values were based on self-complete by study participants, while the DN4-interview validation values were calculated based on a doctor-facilitated interview and assessment, which is not how the tool was used in the epidemiological studies covered in this review. Which instrument performed better in practice is unclear, but it is clear from the difference in the apparent estimates that they performed quite noticeably differently, and that Bayesian correction minimizes the magnitude of the difference in estimates to probably within a reasonable error.

In conclusion, we used a Bayesian methodology to estimate the corrected prevalence of pain with neuropathic characteristics, finding consistent, and substantial downward revisions in prevalence estimates. Our findings may suggest that we need to reappraise how the burden of neuropathic pain is conceptualized in population health. However, even with a global prevalence of 2 to 5% in the adult population, neuropathic pain, with its debilitating effects, still represents a massive burden (∼120 to 300 million people) that requires continued attention.

## Data Availability

All data, analysis scripts, and the analysis environment are available.

https://zenodo.org/records/23695163?preview=1&token=eyJhbGciOiJIUzUxMiJ9.eyJpZCI6ImUzNDRjMzRiLWJmYTItNDMxYS04NTc1LTE4MDczMTkzNmIwNyIsImRhdGEiOnt9LCJyYW5kb20iOiI4NDU5ZjhiN2JlYzk2OGYyNTc2YmNkN2YzYWE1ZmY4ZCJ9.q66R1q0gHTh49UCU2weOLUaLE0dIfunfOUqfjazOmKcV4JG30ToexQBLqhePtfIooJ92CcUmM706mlHXc-uRUg

## Statements and declarations

### Contributions

PRK: Conceptualisation, data curation, formal analysis (including directly accessing and verifying the data), investigation, supervision, visualisation, and writing – original draft. TH: Investigation, and writing – review & editing. NM: Investigation, and writing – review & editing. PCC: Investigation, and writing – review & editing. All authors had full access to all the data and had final responsibility for the decision to submit for publication.

### Competing interests

PRK declares that he is the sole proprietor of Blueprint Analytics and has received honoraria from Partners in Research. TH, NM, and PCC declare no competing interests.

### Data sharing

All data and analysis scripts are available at Zenodo.org: https://zenodo.org/records/23695163?preview=1&token=eyJhbGciOiJIUzUxMiJ9.eyJpZCI6ImUzNDRjMzRiLWJmYTItNDMxYS04NTc1LTE4MDczMTkzNmIwNyIsImRhdGEiOnt9LCJyYW5kb20iOiI4NDU5ZjhiN2JlYzk2OGYyNTc2YmNkN2YzYWE1ZmY4ZCJ9.q66R1q0gHTh49UCU2weOLUaLE0dIfunfOUqfjazOmKcV4JG30ToexQBLqhePtfIooJ92CcUmM706mlHXc-uRUg

### Funding

NM received funding from the National Research Foundation of South Africa. The funder had no input on the project.

## Notes

### Competing Interest Statement

P.R.K. is the sole proprietor of Blueprint Analytics and is a consultant for Partners in Research. The remaining authors have no conflicts of interest to declare.

### Clinical Protocols

https://www.crd.york.ac.uk/PROSPERO/view/CRD42023416845

### Summary of Updates

The revision removes the meta-analysis of prevalence estimates from the eight studies included in the systematic review (study heterogeneity was too high and the number of studies too low to warrant the meta-analysis). The meta-analysis has been replaced by a more comprehensive Bayesian analysis of the data. The initial study used an uninformative prior and a single informative prior. The new corrected prevalences were estimated using Bayesian models that incorporated sensitivity and specificity distributions under four different priors [noninformative prior: Beta(1, 1), informative prior with a mode of 2.5%: Beta(1.398, 16.522), informative prior with a mode of 5%: Beta(2.063, 21.197), and informative prior with a mode of 10%: Beta(5.619, 42.5712)].

