## Supplementary materials for "A systematic review with Bayesian modelling of the prevalence of neuropathic pain"

### Supplementary file

### Section 1

**PRISMA checklist 2020**

| Section and Topic | Ite m # | Checklist item | Location where item is reported |
|---|---|---|---|
| **TITLE** | | | |
| Title | 1 | Identify the report as a systematic review. | 1 |
| **ABSTRACT** | | | |
| Abstract | 2 | See the PRISMA 2020 for Abstracts checklist. | 2 |
| **INTRODUCTION** | | | |
| Rationale | 3 | Describe the rationale for the review in the context of existing knowledge. | 4-5 |
| Objectives | 4 | Provide an explicit statement of the objective(s) or question(s) the review addresses. | 5 |
| **METHODS** | | | |
| Eligibility criteria | 5 | Specify the inclusion and exclusion criteria for the review and how studies were grouped for the syntheses. | 5-6 |
| Information sources | 6 | Specify all databases, registers, websites, organisations, reference lists and other sources searched or consulted to identify studies. Specify the date when each source was last searched or consulted. | 6 |
| Search strategy | 7 | Present the full search strategies for all databases, registers and websites, including any filters and limits used. | Supplement |
| Selection process | 8 | Specify the methods used to decide whether a study met the inclusion criteria of the review, including how many reviewers screened each record and each report retrieved, whether they worked independently, and if applicable, details of automation tools used in the process. | 7 |
| Data collection process | 9 | Specify the methods used to collect data from reports, including how many reviewers collected data from each report, whether they worked independently, any processes for obtaining or confirming data from study investigators, and if applicable, details of automation tools used in the process. | 7-8 |
| Data items | 10a | List and define all outcomes for which data were sought. Specify whether all results that were compatible with each outcome domain in each study were sought (e.g. for all measures, time points, analyses), and if not, the methods used to decide which results to collect. | Supplement |
| | 10b | List and define all other variables for which data were sought (e.g. participant and intervention characteristics, funding sources). Describe any assumptions made about any missing or unclear information. | Supplement |
| Study risk of bias assessment | 11 | Specify the methods used to assess risk of bias in the included studies, including details of the tool(s) used, how many reviewers assessed each study and whether they worked independently, and if applicable, details of automation tools used in the process. | 8 & Supplement |
| Effect measures | 12 | Specify for each outcome the effect measure(s) (e.g. risk ratio, mean difference) used in the synthesis or presentation of results. | 8 |
| Synthesis methods | 13a | Describe the processes used to decide which studies were eligible for each synthesis (e.g. tabulating the study intervention characteristics and comparing against the planned groups for each synthesis (item #5)). | 8-11 |
| | 13b | Describe any methods required to prepare the data for presentation or synthesis, such as handling of missing summary statistics, or data conversions. | 8-11 |
| | 13c | Describe any methods used to tabulate or visually display results of individual studies and syntheses. | 8-11 |
| | 13d | Describe any methods used to synthesize results and provide a rationale for the choice(s). If meta-analysis was performed, describe the model(s), method(s) to identify the presence and extent of statistical heterogeneity, and software package(s) used. | 8-11 |
| | 13e | Describe any methods used to explore possible causes of heterogeneity among study results (e.g. subgroup analysis, meta-regression). | None |
| | 13f | Describe any sensitivity analyses conducted to assess robustness of the synthesized results. | None |
| Reporting bias assessment | 14 | Describe any methods used to assess risk of bias due to missing results in a synthesis (arising from reporting biases). | None |

| Section and Topic | Item # | Checklist item | Location where item is reported |
|---|---|---|---|
| Certainty assessment | 15 | Describe any methods used to assess certainty (or confidence) in the body of evidence for an outcome. | None |
| **RESULTS** | | | |
| Study selection | 16a | Describe the results of the search and selection process, from the number of records identified in the search to the number of studies included in the review, ideally using a flow diagram. | 11-12 & Figure 1 |
| | 16b | Cite studies that might appear to meet the inclusion criteria, but which were excluded, and explain why they were excluded. | 11 & Supplement |
| Study characteristics | 17 | Cite each included study and present its characteristics. | 13-14 & Table 1 |
| Risk of bias in studies | 18 | Present assessments of risk of bias for each included study. | 14 & Table 2 |
| Results of individual studies | 19 | For all outcomes, present, for each study: (a) summary statistics for each group (where appropriate) and (b) an effect estimate and its precision (e.g. confidence/credible interval), ideally using structured tables or plots. | Figures 2-4 |
| Results of syntheses | 20a | For each synthesis, briefly summarise the characteristics and risk of bias among contributing studies. | 12, Figures 2-4 |
| | 20b | Present results of all statistical syntheses conducted. If meta-analysis was done, present for each the summary estimate and its precision (e.g. confidence/credible interval) and measures of statistical heterogeneity. If comparing groups, describe the direction of the effect. | 14-18 |
| | 20c | Present results of all investigations of possible causes of heterogeneity among study results. | None |
| | 20d | Present results of all sensitivity analyses conducted to assess the robustness of the synthesized results. | None |
| Reporting biases | 21 | Present assessments of risk of bias due to missing results (arising from reporting biases) for each synthesis assessed. | None |
| Certainty of evidence | 22 | Present assessments of certainty (or confidence) in the body of evidence for each outcome assessed. | None |
| **DISCUSSION** | | | |
| Discussion | 23a | Provide a general interpretation of the results in the context of other evidence. | 18-19 |
| | 23b | Discuss any limitations of the evidence included in the review. | 19-20 |
| | 23c | Discuss any limitations of the review processes used. | 19-20 |
| | 23d | Discuss implications of the results for practice, policy, and future research. | 22 |
| **OTHER INFORMATION** | | | |
| Registration and protocol | 24a | Provide registration information for the review, including register name and registration number, or state that the review was not registered. | 5 |
| | 24b | Indicate where the review protocol can be accessed, or state that a protocol was not prepared. | 5 |
| | 24c | Describe and explain any amendments to information provided at registration or in the protocol. | NA |
| Support | 25 | Describe sources of financial or non-financial support for the review, and the role of the funders or sponsors in the review. | 24 |
| Competing interests | 26 | Declare any competing interests of review authors. | 23 |
| Availability of data, code and | 27 | Report which of the following are publicly available and where they can be found: template data collection forms; data extracted from included | 8 & 23 |

| Section and Topic | Item # | Checklist item | Location where item is reported |
|---|---|---|---|
| other materials | | studies; data used for all analyses; analytic code; any other materials used in the review. | |

*From:* Page MJ, McKenzie JE, Bossuyt PM, Boutron I, Hoffmann TC, Mulrow CD, et al. The PRISMA 2020 statement: an updated guideline for reporting systematic reviews. BMJ 2021;372:n71. doi: 10.1136/bmj.n71. This work is licensed under CC BY 4.0. To view a copy of this license, visit https://creativecommons.org/licenses/by/4.0/

### Section 2

#### Search strategy

**PubMed**

1: (pain) OR (painful)

2: ((neuropathy) OR (neuropathic)) OR (neuralgia)

3: (((dn4) OR ("douleur neuropathique")) OR ("douleur neuropathique 4")) OR ("douleur neuropathique en 4")

4: (lanss) OR ("leeds assessment of neuropathic symptoms and signs")

5: paindetect

6: ((epidemiology) OR (epidemiological)) OR (prevalence)

7: ("2001/01/01"[Date - Publication] : "2023/03/31"[Date - Publication])

8: Review[Publication Type]

9: #1 AND #2

10: #3 OR #4 OR #5

11: #9 AND #10

12: #11 AND #6

13: #12 AND #7

14: #13 NOT #8

15: #14 Filters: Abstract, Humans, English

**Scopus**

1: TITLE-ABS-KEY ( pain  OR  painful )

2: TITLE-ABS-KEY ( neuropathy  OR  neuropathic  OR  neuralgia )

3: TITLE-ABS-KEY ( dn4  OR  {douleur neuropathique}  OR  {douleur neuropathique 4}  OR  {douleur neuropathique en 4} )

4: TITLE-ABS-KEY ( lanss  OR  {leeds Assessment of neuropathic symptoms and signs} )

5: TITLE-ABS-KEY ( paindetect )

6: TITLE-ABS-KEY ( epidemiology  OR  epidemiological  OR  prevalence )

7: #1 AND #2

8: #3 OR #4 OR #5

9: #7 AND #8

10: #9 AND #6

11: #10 AND  ( EXCLUDE ( DOCTYPE ,  "re" ) )  AND  ( LIMIT-TO ( EXACTKEYWORD ,  "Human" ) )

**Web of Science**

1: (ALL=(pain)) OR ALL=(painful)

2: ((ALL=(neuropathy)) OR ALL=(neuropathic)) OR ALL=(neuralgia)

3: (((ALL=(dn4)) OR ALL=("douleur neuropathique")) OR ALL=("douleur neuropathique 4")) OR ALL=("douleur neuropathique en 4")

4: (ALL=(lanss)) OR ALL=("leeds assessment of neuropathic symptoms and signs")

5: ALL=(paindetect)

6: ((ALL=(epidemiology)) OR ALL=(epidemiological)) OR ALL=(prevalence)

7: #1 AND #2

8: #3 OR #4 OR #5

9: #7 AND #8

10: #9 AND #6 Timespan: 2001-01-01 to 2023-03-31

11: #9 AND #6 and Review Article (Exclude - Document Types) Timespan: 2001-01-01 to 2023-03-31

12: #9 AND #6 and Review Article (Exclude - Document Types) and English (Languages) Timespan: 2001-01-01 to 2023-03-31

### Section 3

#### Data extraction template

(reformatted from the template created in Covidence)

**Country in which the study was conducted**

Your answer

**Region or state in which the study was conducted**

If more than one, list

Your answer

**Region or state in which the study was conducted**

If more than one, list

Your answer

**Characteristics of included studies**

**Aim of the study**

Insert text from paper

Your answer

**Study design**

○ Prevalence study

○ Other: ____________________

**Start date**

YYYY-MM-DD

Your answer ____________________

**End date**

YYYY-MM-DD

Your answer ____________________

**Declaration of conflicts of interest**

○ Declared

○ None declared

**Screening tool used**

- ○ DN4
- ○ DN4 - interview
- ○ LANSS
- ○ S-LANSS
- ○ PainDetect

**Threshold score used to define the presence of neuropathic pain**
List threshold scores if more than one questionnaire is used (e.g., DN4 - interview: ≥ 3, S-LANSS ≥ 12)

Your answer

**Minimum pain duration for classification of chronic pain**

Your answer

**Minimum pain duration for classification of chronic pain**

Your answer

**Participants**

**Inclusion criteria**
If the information is not reported, write, "Not reported".

Your answer

**Exclusion criteria**
If the information is not reported, write, "Not reported".

Your answer

**Sample frame**
If the information is not reported, write, "Not reported".

Your answer

**Method of recruitment of participants**

Choose all that are appropriate.

- [ ] Phone
- [ ] Mail
- [ ] Door-to-door
- [ ] Other: ____________

**Results**

**Total number of people contacted**

Your answer

**Total number of people recruited**

Your answer

**Baseline Population Characteristics**

- If the information is not reported, write, "Not reported".
- If Age (years): add units of measure (e.g., mean = 34, SD = 4)
- If Age (counts): Add counts per category (e.g., 18-25 years = 250)

| | **Overall** | **Acute pain** (any type) | **Chronic pain** (any type) | **Chronic neuropathic pain** | **No pain** |
|---|---|---|---|---|---|
| **Age** (years) | | | | | |
| **Age** (categories) | | | | | |
| **Sex** (% female) | | | | | |

**Prevalence of pain**

Per cent is the percentage of the total sample. If the information is not reported, write, "Not reported".

| | **Count** (n) | **Per cent** (%) | **95% confidence interval for the percentage** |
|---|---|---|---|
| **Respondents with acute pain** (any type) | | | |
| **Respondents with chronic pain** (any type) | | | |
| **Respondents with chronic neuropathic pain** | | | |
| **Respondents with no pain** | | | |

**Intensity of pain**

Convert 0-10 NRS to 0-100 VAS

| | **Intensity** (VAS) | **Intensity** (mild/moderate/severe) |
|---|---|---|
| **Respondents with acute pain** (any type) | | |
| **Respondents with chronic pain** (any type) | | |
| **Respondents with chronic neuropathic pain** | | |

### Section 4

#### Quality assessment template

(reformatted from the template created in Covidence)

**Was the sample frame appropriate to address the target population?**

- ○ Yes
- ○ No
- ○ Unclear

**Were study participants sampled in an appropriate way?**

- ○ Yes
- ○ No
- ○ Unclear

**Was the sample size adequate?**

- ○ Yes
- ○ No
- ○ Unclear

**Were the study subjects and the setting described in detail?**

- ◯ Yes
- ◯ No
- ◯ Unclear

**Was the data analysis conducted with sufficient coverage of the identified sample?**

- ◯ Yes
- ◯ No
- ◯ Unclear

**Were valid methods used for the identification of the condition?**

- ◯ Yes
- ◯ No
- ◯ Unclear

**Was the condition measured in a standard, reliable way for all participants?**

- ○ Yes
- ○ No
- ○ Unclear

**Was there appropriate statistical analysis?**

- ○ Yes
- ○ No
- ○ Unclear

**Was the response rate adequate, and if not, was the low response rate managed appropriately?**

- ○ Yes
- ○ No
- ○ Unclear

### Section 5

#### Beta distributions

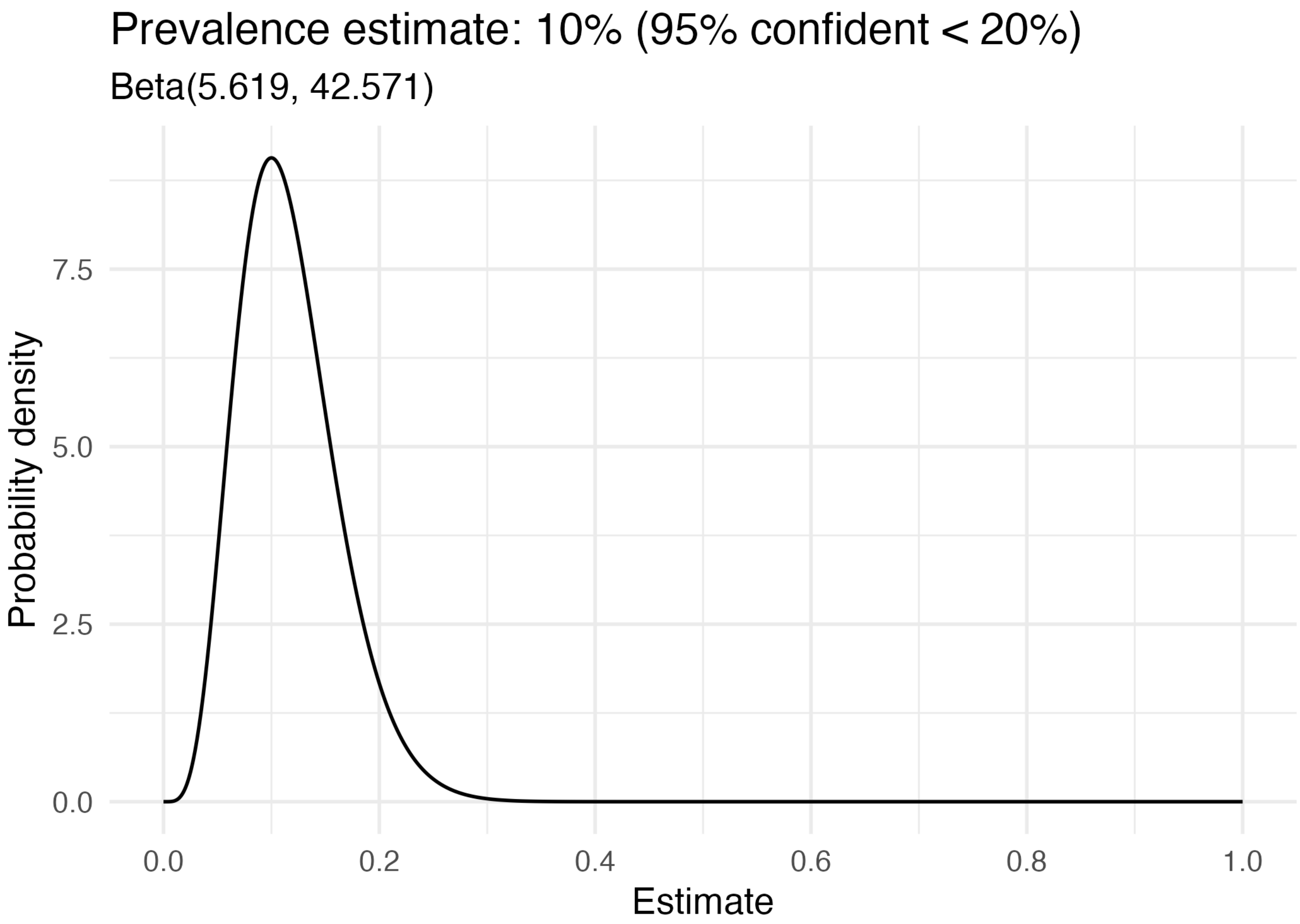
Prevalence estimate: 10% (95% confident < 20%)
Beta(5.619, 42.571)
Probability density
7.5
5.0
2.5
0.0
0.0
0.2
0.4
0.6
0.8
1.0
Estimate

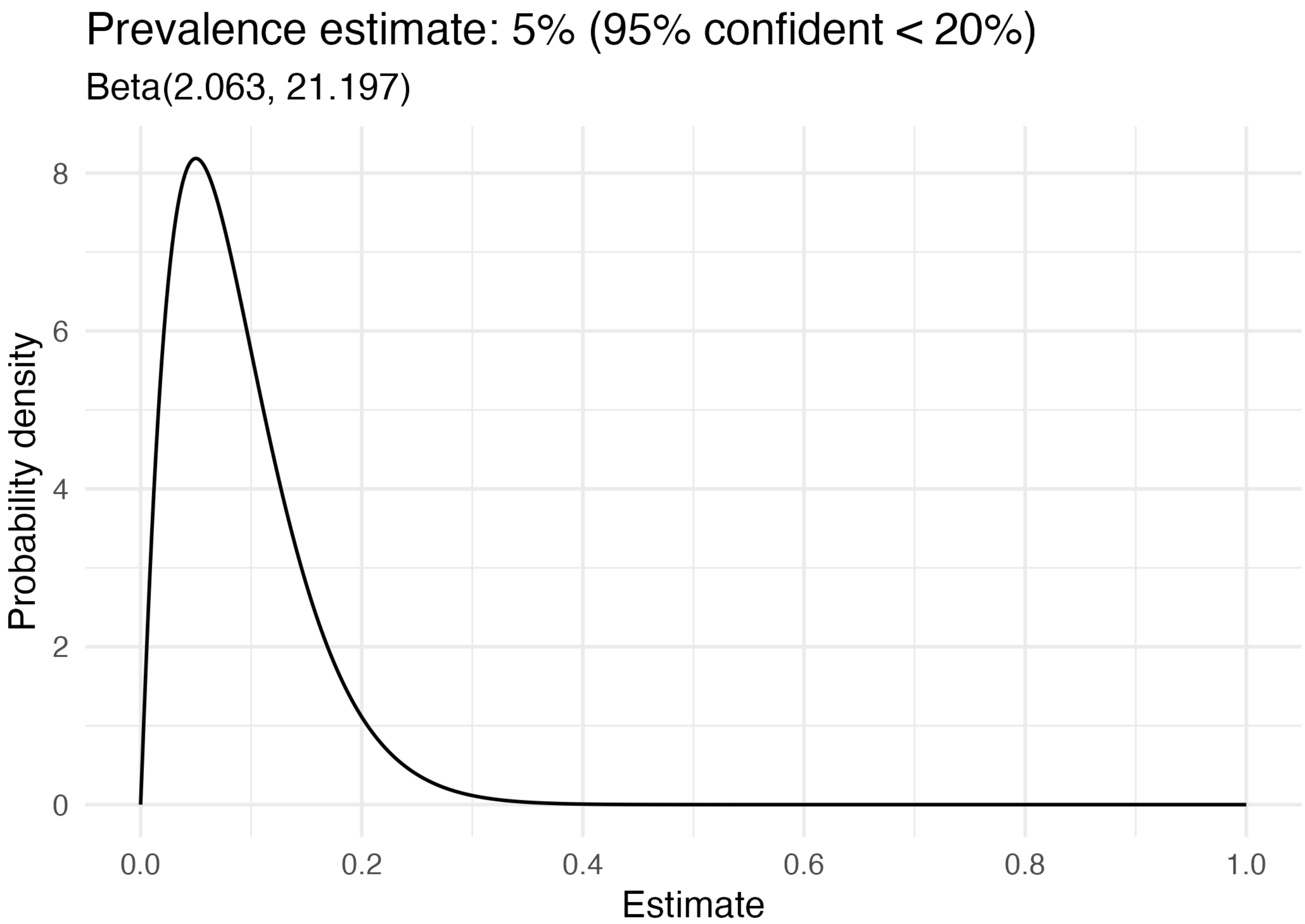
Prevalence estimate: 5% (95% confident < 20%)
Beta(2.063, 21.197)
Probability density
0
2
4
6
8
0.0
0.2
0.4
0.6
0.8
1.0
Estimate

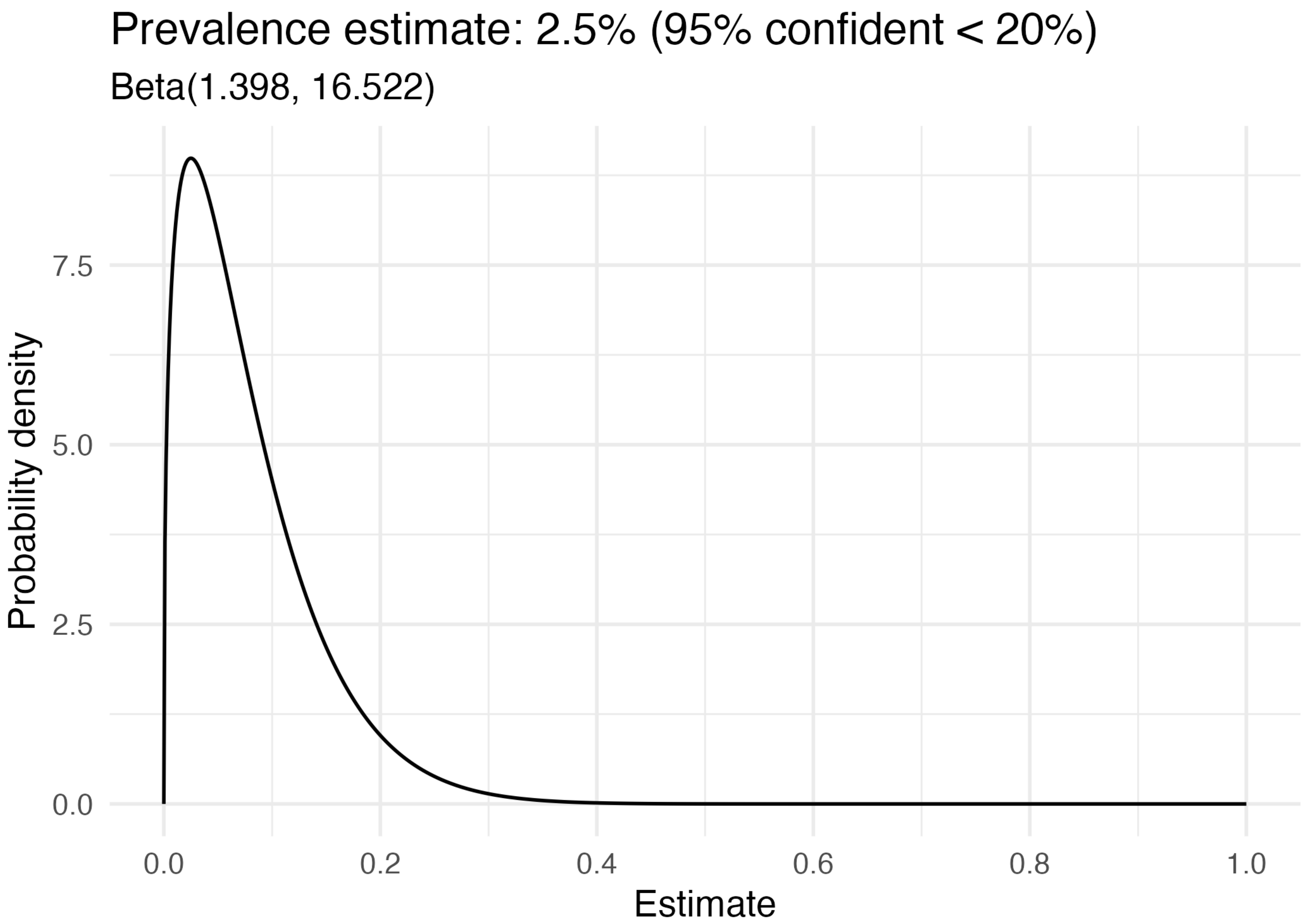
Prevalence estimate: 2.5% (95% confident < 20%)
Beta(1.398, 16.522)
Probability density
7.5
5.0
2.5
0.0
0.0
0.2
0.4
0.6
0.8
1.0
Estimate

### Section 6

#### Excluded studies

**STUDIES EXCLUDED DURING FULL-TEXT SCREENING (N = 12)**

1. Adoukonou T, Gnonlonfoun D, Kpozehouen A, Adjien C, Tchaou B, Tognon-Tchegnonsi F, Adechina H, Covi R, Houinato D. Prévalence et caractéristiques des douleurs chroniques avec caractère neuropathique en population générale à Parakou au nord du Bénin en 2012. Rev. Neurol. (Paris) 2014;170:703–711. doi:10.1016/j.neurol.2014.07.013.

   **Reason for exclusion:** Published in French and a single city (Parakou, Benin)

2. Attal N, Lanteri-Minet M, Laurent B, Fermanian J, Bouhassira D. The specific disease burden of neuropathic pain: results of a French nationwide survey. Pain 2011;152:2836–2843. doi:10.1016/j.pain.2011.09.014.

   **Reason for exclusion:** Sub-study of the publication included in the systematic review and meta-analysis (Bouhassira D, et al. Pain 2008;136:380–387. doi:10.1016/j.pain.2007.08.013), with no additional information being presented on the prevalence of neuropathic pain in the surveyed population.

3. Durán J, Tejos-Bravo M, Cid V, Ferreccio C, Calvo M. Chronic pain in Chile: first prevalence report of noncancer chronic pain, fibromyalgia, and neuropathic pain and its associated factors. Pain 2023;164:1852–1859. doi:10.1097/j.pain.0000000000002886.

   **Reason for exclusion:** Survey of a single city (Molina, Chile).

4. Elzahaf RA, Johnson MI, Tashani OA. The epidemiology of chronic pain in Libya: a cross-sectional telephone survey. BMC Public Health 2016;16:776. doi:10.1186/s12889-016-3349-6.

   **Reason for exclusion:** Survey of three cities (Tripoli, Benghazi, and Sabha; Libya)

5. Elzahaf RA, Tashani OA, Johnson MI. Prevalence of chronic pain among Libyan adults in Derna City: a pilot study to assess the reliability, linguistic validity, and feasibility of using an Arabic version of the structured telephone interviews questionnaire on chronic pain. Pain Pract. 2013;13:380–389. doi:10.1111/j.1533-2500.2012.00594.x.

   **Reason for exclusion:** Survey of a single city (Derna City, Libya).

6. Hamdan A, Luna JD, Del Pozo E, Gálvez R. Diagnostic accuracy of two questionnaires for the detection of neuropathic pain in the Spanish population: Comparison of neuropathic pain questionnaires. Eur. J. Pain 2014;18:101–109. doi:10.1002/j.1532-2149.2013.00350.x.

   **Reason for exclusion:** Assessed a patient population at two chronic pain clinics.

7. Hébert HL, Veluchamy A, Baskozos G, Fardo F, Van Ryckeghem D, Pearson ER, Colvin LA, Crombez G, Bennett DLH, Meng W, Palmer CNA, Smith BH. Development and external validation of multivariable risk models to predict incident and resolved neuropathic pain: a DOLORisk Dundee study. J. Neurol. 2023;270:1076–1094. doi:10.1007/s00415-022-11478-0.

   **Reason for exclusion: French language publication**

8. de Moraes Vieira EB, Garcia JBS, da Silva AAM, Mualem Araújo RLT, Jansen RCS. Prevalence, characteristics, and factors associated with chronic pain with and without neuropathic characteristics in são Luís, Brazil. J. Pain Symptom Manage. 2012;44:239–251. doi:10.1016/j.jpainsymman.2011.08.014.

   **Reason for exclusion:** Survey of a single municipality (São Luís, Brazil)

9. Smith BH, Torrance N, Bennett MI, Lee AJ. Health and quality of life associated with chronic pain of predominantly neuropathic origin in the community. Clin. J. Pain 2007;23:143–149. doi:10.1097/01.ajp.0000210956.31997.89.

   **Reason for exclusion:** Not a prevalence study; reported on the impact of neuropathic pain using data published in Torrance N, et al. The epidemiology of chronic pain of predominantly neuropathic origin. Results from a general population survey. J Pain. 2006;7:281–289. doi:10.1016/j.jpain.2005.11.008 (see paper 11 below).

10. Torrance N, Ferguson JA, Afolabi E, Bennett MI, Serpell MG, Dunn KM, Smith BH. Neuropathic pain in the community: more under-treated than refractory? Pain 2013;154:690–699. doi:10.1016/j.pain.2012.12.022.

    **Reason for exclusion:** Survey of 10 GP practices in 5 UK locations. Three cities in England: Leeds, Lancaster, and Stafford; one city in Scotland: Glasgow; and one region in Scotland: Grampian. Regional Grampian data were not reported separately.

11. Torrance N, Smith BH, Bennett MI, Lee AJ. The epidemiology of chronic pain of predominantly neuropathic origin. Results from a general population survey. J. Pain 2006;7:281–289. doi:10.1016/j.jpain.2005.11.008.

    **Reason for exclusion:** Survey of two cities (Leeds and London, England) and one region (Grampian, Scotland). Regional Grampian data were not reported separately.

12. Torrance N, Smith BH, Watson MC, Bennett MI. Medication and treatment use in primary care patients with chronic pain of predominantly neuropathic origin. Fam. Pract. 2007;24:481–485. doi:10.1093/fampra/cmm042.

    **Reason for exclusion:** Not a prevalence study; reported on the management of neuropathic pain using data published in Torrance N, et al. The epidemiology of chronic pain of predominantly neuropathic origin. Results from a general population survey. J Pain. 2006;7:281–289. doi:10.1016/j.jpain.2005.11.008 (see paper 11 above).

**STUDY EXCLUDED DURING DATA EXTRACTION (N = 1)**

1. Inoue S, Taguchi T, Yamashita T, Nakamura M, Ushida T. The prevalence and impact of chronic neuropathic pain on daily and social life: A nationwide study in a Japanese population. Eur. J. Pain 2017;21:727–737. doi:10.1002/ejp.977.

    **Reason for exclusion:** Data on the sensitivity and specificity of the Japanese version of painDETECT, when using a threshold score of ≥ 13, were not available. The authors were approached via email, but only information on the specificity was forthcoming.

### Section 7

#### Posterior estimates

(stratified by questionnaire)

#### Table S1. DN4: Douleur Neuropathique en 4 Questions (interview)

| Study | Prior | Prevalence [%; median (95% high-density interval) of the posterior distribution; *point estimate (95% confidence interval)] | Absolute difference between posterior median and apparent prevalence point estimate (% points) | Percentage difference between posterior median and apparent prevalence point estimate (%) | Width of 95% high density interval (% points) |
|---|---|---|---|---|---|
| VanDenKerkhof (2016) | **Apparent** | **11.5 (11.4 to 11.5)*** | | | |
| | Beta(1, 1) | 2 (0 to 6.6) | -9.5 | -82.7 | 6.6 |
| | Beta(1.398, 16.522); mode = 2.5% | 2 (0 to 5.2) | -9.5 | -82.7 | 5.2 |
| | Beta(2.063, 21.197); mode = 5% | 3.2 (0.3 to 7) | -8.3 | -72.1 | 6.7 |
| | Beta(5.619, 42.5712); mode = 10% | 5.9 (2.4 to 9.8) | -5.6 | -48.7 | 7.4 |
| Toth (2009) | **Apparent** | **17.9 (15.8 to 20.2)*** | | | |
| | Beta(1, 1) | 5.3 (0 to 13.9) | -12.6 | -70.4 | 13.9 |
| | Beta(1.398, 16.522); mode = 2.5% | 4.2 (0 to 11.3) | -13.7 | -76.6 | 11.3 |
| | Beta(2.063, 21.197); mode = 5% | 5.4 (0.4 to 12.3) | -12.5 | -69.7 | 11.9 |
| | Beta(5.619, 42.5712); mode = 10% | 8.5 (3.1 to 14.6) | -9.4 | -52.3 | 11.5 |
| Hebert (2021) | **Apparent** | **15.4 (14.5 to 16.3)*** | | | |
| | Beta(1, 1) | 3.8 (0 to 10.7) | -11.6 | -75.6 | 10.7 |
| | Beta(1.398, 16.522); mode = 2.5% | 3.4 (0 to 9.2) | -12.0 | -78.1 | 9.2 |
| | Beta(2.063, 21.197); mode = 5% | 4.5 (0.3 to 10.3) | -10.9 | -70.8 | 10 |
| | Beta(5.619, 42.5712); mode = 10% | 7.5 (2.8 to 12.8) | -7.9 | -51.5 | 10 |

| | | | | | |
|---|---|---|---|---|---|
| Harifi (2013) | **Apparent** | **10.6 (9.8 to 11.5)*** | | | |
| | Beta(1, 1) | 1 (0 to 3.6) | -9.6 | -90.6 | 3.6 |
| | Beta(1.398, 16.522); mode = 2.5% | 1.3 (0 to 3.8) | -9.3 | -88.2 | 3.8 |
| | Beta(2.063, 21.197); mode = 5% | 1.8 (0.1 to 4.5) | -8.8 | -82.9 | 4.4 |
| | Beta(5.619, 42.5712); mode = 10% | 3.8 (1.3 to 6.6) | -6.8 | -64.4 | 5.3 |
| Bouhassira (2008) | **Apparent** | **6.9 (6.6 to 7.2)*** | | | |
| | Beta(1, 1) | 1.1 (0 to 3.6) | -5.8 | -84.7 | 3.6 |
| | Beta(1.398, 16.522); mode = 2.5% | 1.3 (0 to 3.7) | -5.6 | -80.9 | 3.7 |
| | Beta(2.063, 21.197); mode = 5% | 1.8 (0.1 to 4.3) | -5.1 | -73.6 | 4.2 |
| | Beta(5.619, 42.5712); mode = 10% | 3.6 (1.4 to 6.1) | -3.3 | -47.6 | 4.7 |
| Baskozos (2023) | **Apparent** | **9.2 (9.1 to 9.4)*** | | | |
| | Beta(1, 1) | 1.5 (0 to 4.9) | -7.7 | -83.9 | 4.9 |
| | Beta(1.398, 16.522); mode = 2.5% | 1.9 (0 to 5.2) | -7.4 | -79.8 | 5.2 |
| | Beta(2.063, 21.197); mode = 5% | 2.6 (0.2 to 5.9) | -6.7 | -72.3 | 5.7 |
| | Beta(5.619, 42.5712); mode = 10% | 4.9 (1.8 to 8.2) | -4.4 | -47.5 | 6.4 |

#### Table S2. S-LANSS: Self-Administered Leeds Assessment of Neuropathic Symptoms and Signs

| Study | prior | Prevalence [%; median (95% high-density interval) of the posterior distribution; *point estimate (95% confidence interval)] | Absolute difference between posterior median and apparent prevalence point estimate (% points) | Percentage difference between posterior median and apparent prevalence point estimate (%) | Width of 95% high density interval (% points) |
|---|---|---|---|---|---|
| Yawn (2009) | **Apparent** | **8.8 (7.9 to 9.8)*** | | | |
| | Beta(1, 1) | 1.2 (0 to 4.2) | -7.7 | -86.9 | 4.2 |
| | Beta(1.398, 16.522); mode = 2.5% | 1.4 (0 to 4.2) | -7.4 | -84.1 | 4.2 |
| | Beta(2.063, 21.197); mode = 5% | 2 (0.1 to 5) | -6.8 | -77.0 | 4.9 |
| | Beta(5.619, 42.5712); mode = 10% | 4.2 (1.5 to 7.4) | -4.6 | -52.5 | 5.9 |
| VanDenKerkhof (2016) | **Apparent** | **7.7 (7.7 to 7.7)*** | | | |
| | Beta(1, 1) | 0.9 (0 to 3.4) | -6.8 | -88.2 | 3.4 |
| | Beta(1.398, 16.522); mode = 2.5% | 1.2 (0 to 3.6) | -6.5 | -84.1 | 3.6 |
| | Beta(2.063, 21.197); mode = 5% | 1.7 (0.1 to 4.2) | -6.0 | -78.0 | 4.1 |
| | Beta(5.619, 42.5712); mode = 10% | 3.6 (1.3 to 6.2) | -4.1 | -53.5 | 4.9 |

#### Table S3. PainDETECT

| Study | Prior | Prevalence [%; median (95% high-density interval) of the posterior distribution; *point estimate (95% confidence interval)] | Absolute difference between posterior median and apparent prevalence point estimate (% points) | Percentage difference between posterior median and apparent prevalence point estimate (%) | Width of 95% high density interval (% points) |
|---|---|---|---|---|---|
| DiBonaventura (2017) | **Apparent** | **10 (10 to 10)*** | | | |
| | Beta(1, 1) | 1.6 (0 to 5.6) | -8.4 | -84.0 | 5.6 |
| | Beta(1.398, 16.522); mode = 2.5% | 2.4 (0 to 5.6) | -7.7 | -76.5 | 5.6 |
| | Beta(2.063, 21.197); mode = 5% | 3.4 (0.2 to 6.8) | -6.6 | -66.1 | 6.6 |
| | Beta(5.619, 42.5712); mode = 10% | 5.2 (1.8 to 9.1) | -4.8 | -47.7 | 7.3 |
